# Antithrombotic Therapy in COVID-19: Systematic Summary of Ongoing or Completed Randomized Trials

**DOI:** 10.1101/2021.01.04.21249227

**Authors:** Azita H. Talasaz, Parham Sadeghipour, Hessam Kakavand, Maryam Aghakouchakzadeh, Elaheh Kordzadeh-Kermani, Benjamin W. Van Tassell, Azin Gheymati, Hamid Ariannejad, Seyed Hossein Hosseini, Sepehr Jamalkhani, Michelle Sholzberg, Manuel Monreal, David Jimenez, Gregory Piazza, Sahil A. Parikh, Ajay Kirtane, John W. Eikelboom, Jean M. Connors, Beverley J. Hunt, Stavros V. Konstantinides, Mary Cushman, Jeffrey I. Weitz, Gregg W. Stone, Harlan M. Krumholz, Gregory Y.H. Lip, Samuel Z. Goldhaber, Behnood Bikdeli

## Abstract

Endothelial injury and microvascular/macrovascular thrombosis are common pathophysiologic features of coronavirus disease-2019 (COVID-19). However, the optimal thromboprophylactic regimens remain unknown across the spectrum of illness severity of COVID-19. A variety of antithrombotic agents, doses and durations of therapy are being assessed in ongoing randomized controlled trials (RCTs) that focus on outpatients, hospitalized patients in medical wards, and critically-ill patients with COVID-19. This manuscript provides a perspective of the ongoing or completed RCTs related to antithrombotic strategies used in COVID-19, the opportunities and challenges for the clinical trial enterprise, and areas of existing knowledge, as well as data gaps that may motivate the design of future RCTs.

## INTRODUCTION

### Thromboembolism in Patients with COVID-19

Microvascular and macrovascular thrombotic complications including arterial and especially venous thromboembolism (VTE) appear to be common clinical manifestations of coronavirus disease 2019 (COVID-19), particularly among hospitalized and critically-ill patients (1–4). Pooled analyses have helped in providing aggregate estimates of thrombotic events (4, 5). In a recent systematic review and meta-analysis, the overall incidence of VTE among inpatients with COVID-19 was estimated at 17% (95% confidence interval [CI], 13.4%-20.9%), with variation based on study design and method of ascertainment, with a four-fold higher incidence rate in patients in the intensive care units (ICU) compared with non-ICU settings (28% vs. 7%) (6). In addition, post-mortem studies show frequent evidence of microvascular thrombosis in patients with COVID-19 (7, 8). The influence of these events on mortality rates remains unknown (9).

### Pathophysiology of Thromboembolism in COVID-19: Virchow’s Triad in Action

COVID-19 can potentiate all three components of Virchow’s triad and increase the risk of thrombosis (Figure 1). First, SARS-CoV-2 infection may trigger endothelial dysfunction. Using the angiotensin converting enzyme (ACE)-2, expressed on the surface of many cells, SARS-CoV-2 enters endothelial cells and may impair their intrinsic antithrombotic properties. It is proposed that viremia, hypoxia, the inflammatory response, increased expression of tissue factor, and elevated levels of neutrophil extracellular traps (NETs) can together disrupt the hemostasis equilibrium, and promote endothelial activation (10–12). This induction of a procoagulant state along with the reduction in plasminogen activators, further results in increased platelet reactivity (13–15). Inflammatory cytokines and endothelial activation can lead to downregulation of antithrombin and protein C expression, and an increase in the levels of plasminogen activator inhibitor, fibrinogen, factors V, VII, VIII, and X, in addition to von Willebrand factor (16). Increased platelet reactivity, NETosis and alterations in the aforementioned hemostatic factors result in a hypercoagulable state (17–22). Particularly in COVID-19, it is thought that the excessive inflammatory response plays an important role in the pathogenesis of thrombosis (thromboinflammation), including pulmonary microthrombosis and pulmonary intravascular coagulopathy (7, 8). Antiphospholipid antibodies have been identified in some patients (23) but their clinical significance is uncertain (24). Finally, venous stasis may synergistically combine with this prothrombotic milieu in patients more severely affected by the clinical syndrome of COVID-19 constituting of extreme fatigue with decreased daily activity, immobility due to oxygen requirements in the hospital. This may be further exacerbated by development of complications such as viral myocarditis and left ventricular systolic dysfunction (25, 26). All the aforementioned mechanisms may increase the risk of arterial and venous thrombosis, thereby impacting the severity of illness.

**Figure 1.**
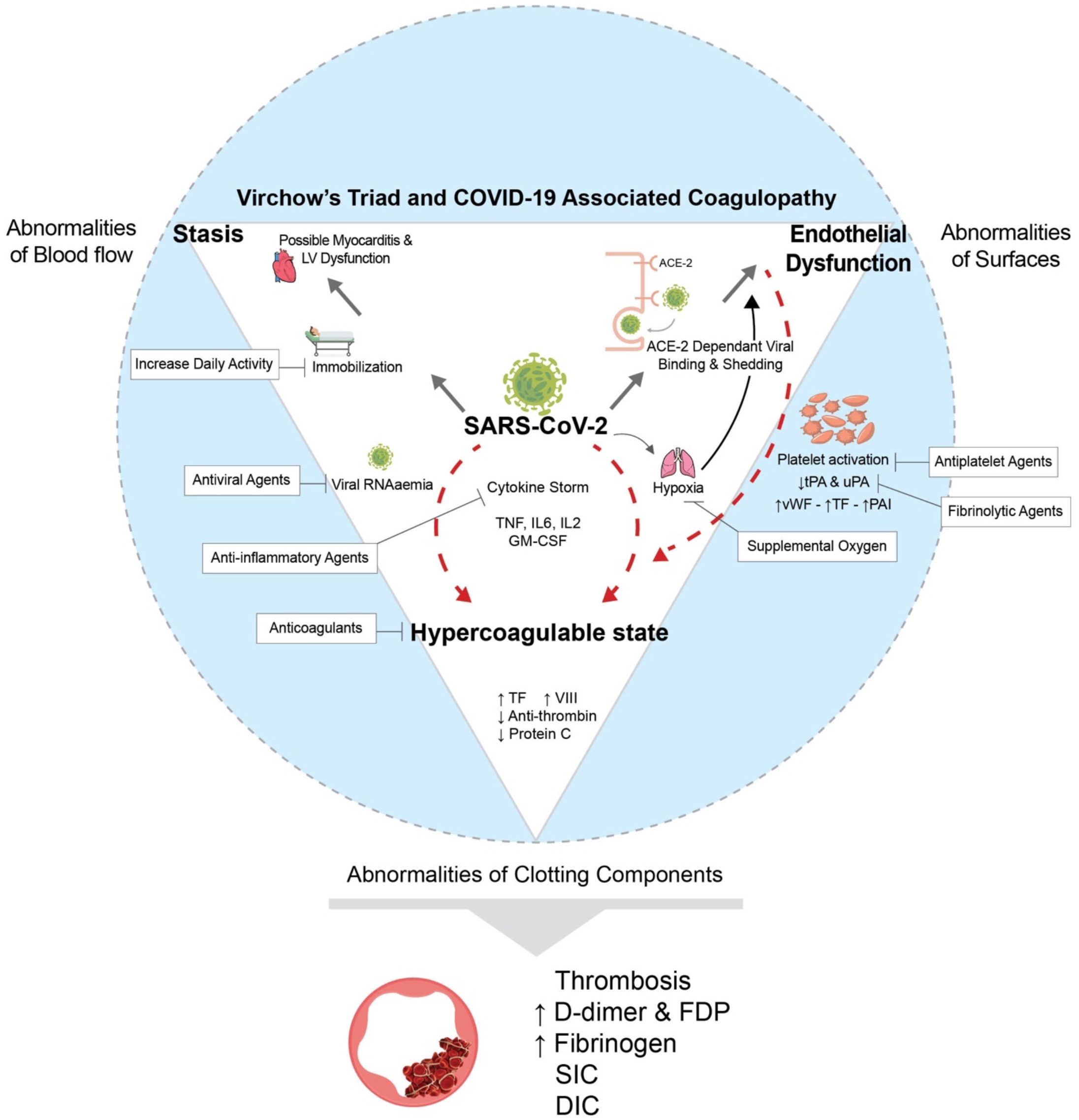
Virchow’s Triad and COVID-19 Associated Coagulopathy. Caption: SARS-CoV-2 can potentiate all three sides of Virchow’s triad including endothelial dysfunction, blood flow stasis, and hypercoagulability. ACE-2 dependent viral entry and the virus-induced inflammatory response can lead to endothelial dysfunction; Bedridden status may lead to stasis; and inflammation, viremia, and cytokine storm can produce a hypercoagulable state. Factor Xa may play a role in spike protein cleavage and endocytosis of the virus. ACE: Angiotensin Converting Enzyme; COVID: Coronavirus Disease 2019; DIC: Disseminated Intravascular Coagulopathy; FDP: Fibrin Degradation Products; GM-CSF: Granulocyte-macrophage colony-stimulating factor; HF: Heart Failure; IL: Interleukin; LV: Left Ventricular; PAI: plasminogen activator inhibitor; RAAS: Renin Angiotensin Aldosterone System; RNA: Ribonucleic Acid; SIC: Sepsis Induced Coagulopathy; TF: Tissue Factor; TNF: Tumor Necrosis Factor; tPA: tissue type plasminogen activators; uPA: urokinase plasminogen activators; vWF: von Willebrand factor.

### Antithrombotic Prophylaxis in COVID-19: Pros and Cons

Bedside observations, pathophysiological investigations, and initial epidemiological data led to enthusiasm for antithrombotic prophylaxis in COVID-19 (27–31). The concern for thrombotic risk was heightened by reports of VTE in 13% to 56% of patients despite the use of standard prophylaxis. (32–35) This led some experts to recommend empiric use of escalated doses of anticoagulants (36). However, the risks associated with intensified use of antithrombotic agents such as bleeding should be weighed against the presumptive benefits (22, 27, 31).

In addition, there have been variations in methodology and outcomes assessment for thrombotic events –including the concern about counting situ thrombosis in small vessels, a recognized feature of acute lung injury also known as immunothrombosis, as pulmonary emboli. Due to these issues, as well as well as the concerns for excess bleeding, a number of guidance statements have not recommended empiric escalated-dose anticoagulation (27, 37).

Multiple ongoing randomized controlled trials (RCTs) are evaluating a variety of antithrombotic regimens in patients with COVID-19 (Figure 2). These include trials of antiplatelet agents, anticoagulants, fibrinolytic agents, or combinations of these agents. In most trials, the intensity of antithrombotic therapy is proportional to the expected thrombotic event rates in the population under study. Less intensive therapies including antiplatelet agents, oral anticoagulants, and standard prophylactic dose of low molecular weight heparin (LMWH) are typically studied in the outpatient or lower acuity hospital settings. In turn, more intensive therapies including intermediate-dose or fully-therapeutic doses of anticoagulants, or even fibrinolytic therapy are under investigation in RCTs of hospitalized critically-ill patients.

**Figure 2.**
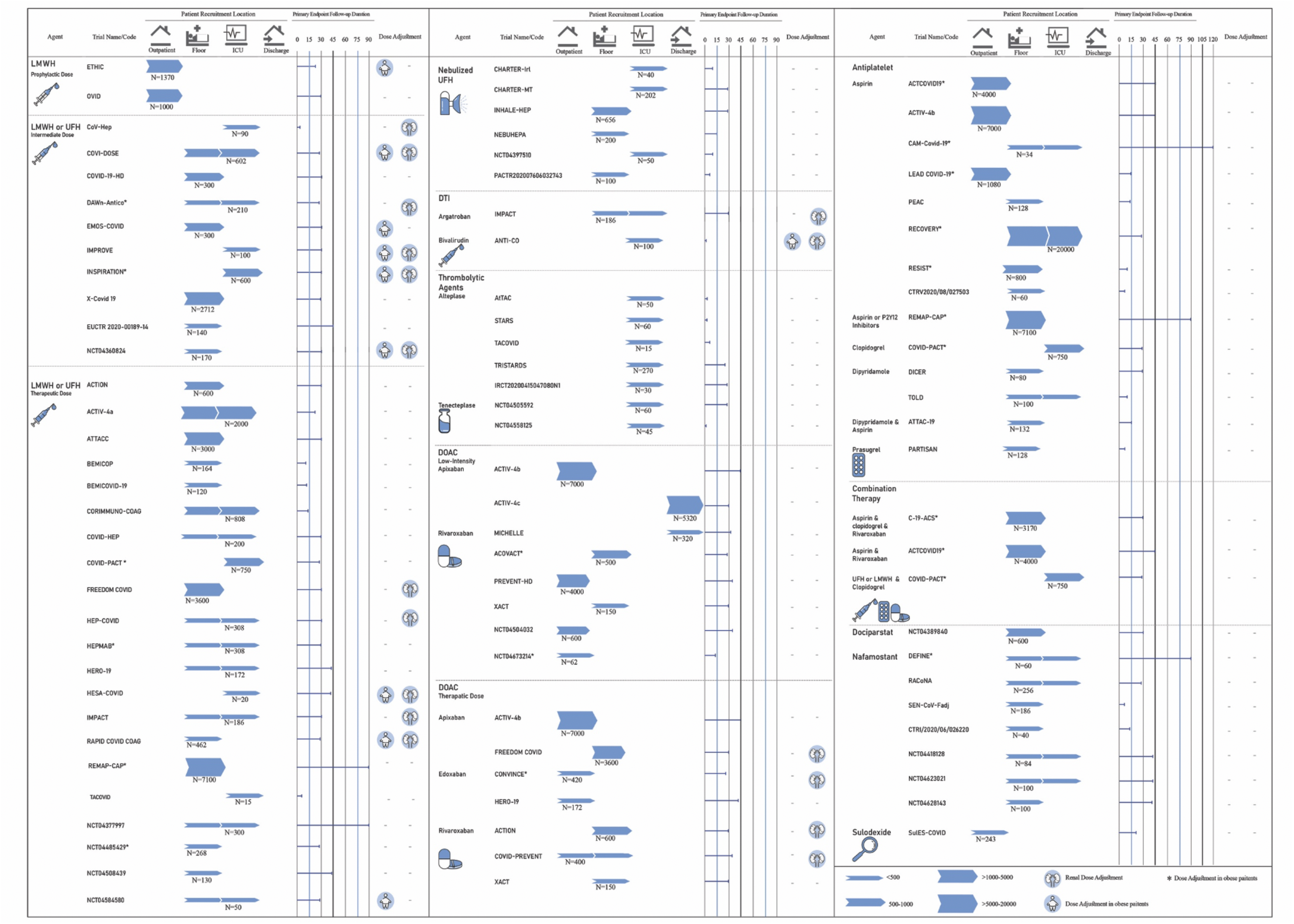
Summary of randomized controlled trials of antithrombotic agents in COVID-19 categorized based on pharmacologic class. Caption: UFH, LMWH, DTI, DOACs, antiplatelets, fibrinolytic, and investigational agents are evaluating in different settings including outpatients, inpatients (ICU and non-ICU), and post-discharge. *: Multifactorial designs or multiple interventions. DOAC: Direct Oral Anticoagulant; DTI: Direct Thrombin Inhibitors; ICU: Intensive Care Unit; LMWH: Low Molecular Weight Heparin; UFH: Unfractionated Heparin.

The aims of this article are to systematically summarize the ongoing and completed RCTs of antithrombotic therapy in patients with COVID-19, to evaluate the strengths and limitations of the study designs, as well as the challenges and opportunities related to conducting and interpreting RCTs during a global pandemic.

## METHODS

We conducted a systematic literature search of trials in clinicaltrials.gov and World Health Organization International Clinical Trials Registry Platform (WHO ICTRP), with the pre-defined keywords of COVID-19, and search terms for antiplatelet agents, anticoagulants, anticoagulation, fibrinolytic agents, and antithrombotic agents. We screened the identified studies and included those that were designed as RCTs with at least one active arm of antithrombotic therapy (date of last search: December 16, 2020). Supplemental Table 1 summarizes study-level inclusion and exclusion criteria for this review.

For the included studies, we searched PubMed and MedRxiv for design papers, study protocols or published results of those studies. The list was complemented by hand-searching and discussion within the author group.

After identification of 918 records and manual screening of 180 records, we included 75 RCTs (Supplemental figure 1). In 13 cases, a design paper and/or study protocol was available. Of all ongoing studies, one RCT reported the results in peer-reviewed literature (38) and one shared the findings on a pre-print server (39). For three RCTs, final results are unknown but patient enrollment was paused in critically-ill patients due to concern for futility and potential excess of safety events (40).

## REVIEW OF ONGOING OR COMPLETED RANDOMIZED CONTROLLED TRIALS

As of December 16, 2020, 75 RCTs of antithrombotic agents for patients with COVID-19 were registered at ClinicalTrials.gov or WHO ICTRP databases. Figure 2 provides a graphical summary of all RCTs of antithrombotic agents in COVID-19 in a pharmacologic-based approach. Agents used in these trials include antiplatelet agents, unfractionated heparin (UFH) and heparin derivatives, parenteral direct thrombin inhibitors (DTIs), direct oral anticoagulants (DOACs), fibrinolytic agents, sulodexide (a mixture of heparan sulfate and dermatan sulfate) (39), dociparstat (a heparin derivative with anti-inflammatory properties), and nafamostat (a synthetic serine protease inhibitor with anticoagulant activity). A succinct discussion of the design features of these trials is provided below according to the clinical setting (see Supplemental Tables 2 and 3 for more details). Supplemental Table 4 summarizes the RCTs of other investigational agents with antithrombotic properties but not meeting our eligibility criteria for the current manuscript.

In each section, the discussion starts with antiplatelet agents, followed by oral anticoagulants, parenteral anticoagulants, fibrinolytic therapy, and investigational agents with antithrombotic properties. This sequence is arbitrary and does not indicate treatment preference. Figure 3 illustrates how RCTs of various agents can fill the knowledge gaps about antithrombotic therapy in COVID-19 in various settings of illness severity.

**Figure 3.**
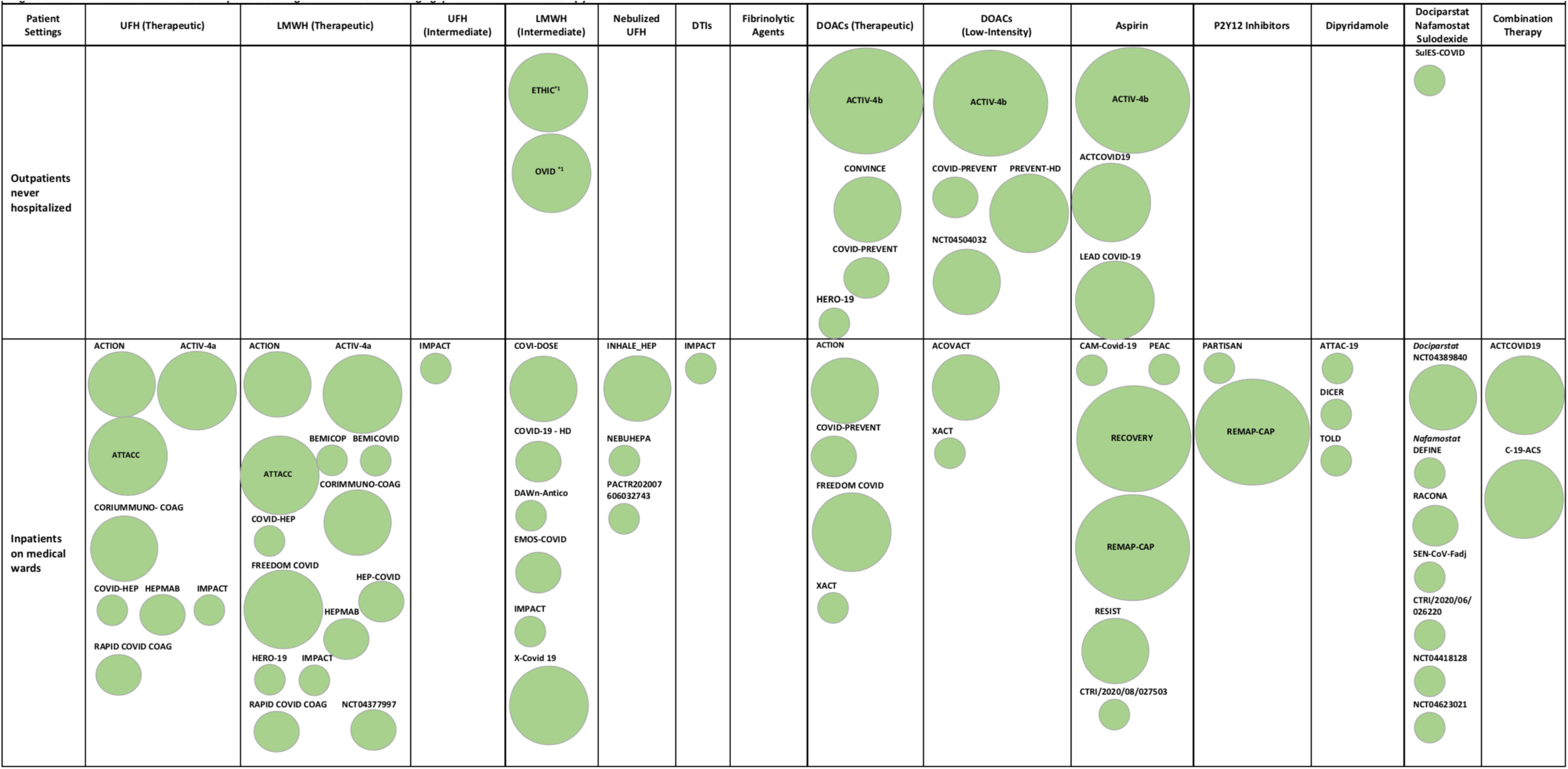
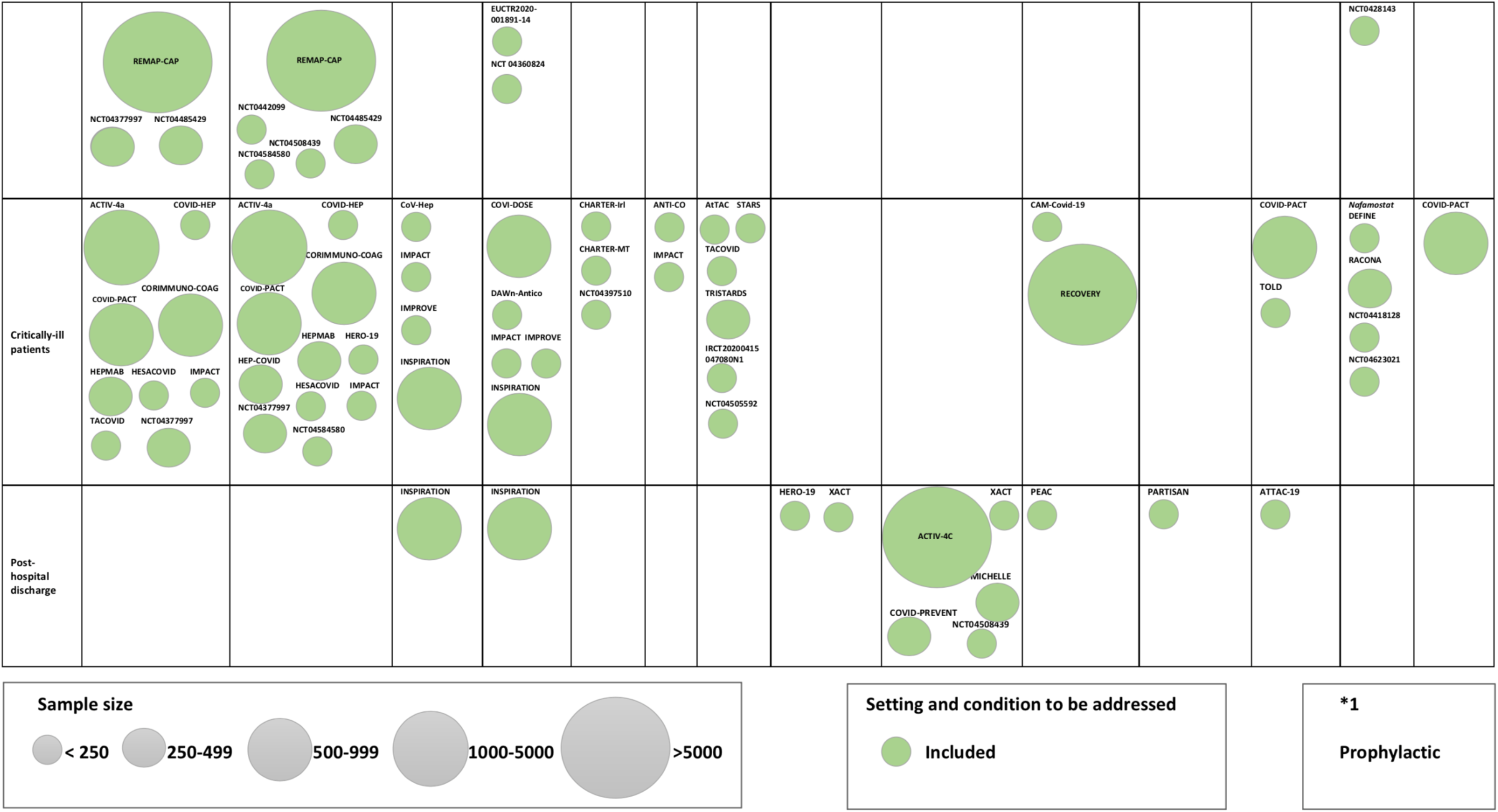
Graphical Summary of ongoing RCTs of antithrombotic therapy in COVID-19 based on patient settings. Caption: Categorizing the RCTs evaluating different agents in different settings including those treated entirely as outpatient, patients in the non-ICU hospital wards, critically ill patients in the ICU, and post-hospital discharge. Others: dociparstat, nafamostat, and sulodexide. DOAC: Direct Oral Anticoagulants; DTI: Direct Thrombin Inhibitors; LMWH: Low Molecular Weight Heparin; RCT: Randomized Clinical Trials; UFH: Unfractionated Heparin.

### Ongoing Clinical Trials of Antithrombotic Agents in the Outpatient Setting

Eleven RCTs of antithrombotic therapy in outpatients with COVID-19 have been registered in clinical trials databases and are studying aspirin, DOACs, enoxaparin, and sulodexide compared with no treatment (6/11) or with placebo (5/11). These trials are mostly (8/11) open-label, with the number of participants ranging from 172 to 7,000 patients, and include patients with a hyperinflammatory or procoagulant profile (including elevated levels of C-reactive protein [CRP] (1/11) or d-dimer [2/11]) and exclude patients at high risk of bleeding (such as those with history of recent gastrointestinal bleeding or intracranial hemorrhage). Pregnant women and patients with severe kidney dysfunction (creatinine clearance [CrCl] < 30 mL/min) are excluded from 8 and 6 of these trials, respectively. The most common primary outcome in the outpatient trials include the need for hospitalization, incidence of thromboembolic events, mortality or composite outcomes inclusive of these within 21 to 90 days after randomization.

Aspirin, DOACs (at both low-intensity and high-intensity), LMWHs (at standard prophylactic dose), and sulodexide are the agents under investigation in the outpatient setting. The impact of low-dose aspirin on the composite rate of hospitalizations and mortality is being evaluated in 3 RCTs with a total of 12,080 patients with COVID-19 (ACTCOVID19, LEAD COVID-19, and ACTIV-4b). Low-intensity rivaroxaban (10 mg QD) is being evaluated in a total of 4,600 patients in two ongoing RCTs (PREVENT-HD [NCT04508023] and NCT04504032). Low-intensity apixaban (2.5 mg BID) is also under investigation in the ACTIV-4b trial in up to 7,000 patients. High-intensity DOACs including rivaroxaban (20 mg QD), apixaban (5 mg BID), and edoxaban (60 mg QD) are being investigated among 7,992 patients in 4 RCTs (COVID-PREVENT, ACTIV-4b, HERO-19, and CONVINCE). The primary outcome for the COVID-PREVENT, HERO-19 and CONVINCE trials is the composite of mortality and arterial and venous thromboembolism; the primary outcome for the randomized double-blind placebo controlled ACTIV-4b trial is a composite of venous and arterial thromboembolism, hospitalization for cardiovascular/pulmonary events and all-cause mortality. ETHIC and OVID RCTs are comparing the effect of standard prophylactic dose of enoxaparin versus no intervention on the primary outcome of hospitalization or mortality in 2,370 individuals (41).

SulES-COVID is the only completed trial of antithrombotic therapy in outpatients with COVID-19. This single-center study of 243 participants assessed the efficacy of sulodexide, compared with placebo on 21-day rates of hospitalization and need for use of supplemental oxygen. Use of sulodexide was associated with reduced hospital admissions (relative risk [RR], 0.6; 95% CI, 0.37-0.96; p=0.03) and need for oxygen support (RR, 0.71; 95% CI, 0.5 to 1; p=0.05), without a significant effect on mortality (39). The study has limitations, including frequent (22.1%) post-enrollment exclusions due to negative SARS-CoV2 test results or lost to follow-up.

Many of the outpatient antithrombotic therapy trials for COVID-19 are large and the follow-up windows are sufficient to capture the intended primary outcomes. An issue with some of these trials is an open-label design, which is a pragmatic feature facilitating the design and enrollment but potentially limits the internal validity, especially for outcomes that may be less bias-resistant. In addition, the available data do not clarify whether dose adjustments are made for renal or liver dysfunction.

### Ongoing Clinical trials of Antithrombotic Agents in Hospitalized non-ICU Patients

We identified 50 ongoing RCTs related to antithrombotic therapy in hospitalized non-ICU patients with COVID-19. Most trials (44/50) are open-label. The antithrombotic agents under investigation include aspirin, P2Y12 inhibitors, dipyridamole, DOACs, heparin (both systemic and inhaled), dociparstat, nafamostat, and a combination of these drugs. The planned sample sizes range between 34 and 20,000 patients. Considering the potential link between elevated D-dimer, micro and macro-thrombosis in COVID-19, and worse outcomes in COVID-19 (42–44), many RCTs (N=16) include patients with elevated D-dimer levels with cut-offs ranging from >500 ng/mL to >1,500 ng/mL (or defined as >2-4 times the upper limit of normal per the local laboratory). Most trials exclude pregnant women (41 studies) and patients with active bleeding or history of intracranial or gastrointestinal bleeding (39 studies). Many trials also excluded patients with CrCl < 30 mL/min (20 studies). In most trials, the time frame for the primary outcome assessment is 28-30 days, although a few studies are designed to assess the primary outcomes at earlier or longer durations. These RCTs are focused on primary efficacy outcomes including all-cause mortality, VTE, arterial thrombosis, requirement for respiratory support, or a composite of these outcomes.

The potential protective effect of antiplatelet agents in hospitalized patients with COVID-19 is being evaluated in 11 RCTs. REMAP-CAP is a large global RCT with a multifactorial adaptive design which is planning to randomize 7,100 patients to multiple therapeutic interventions including an anticoagulant arm and antiplatelet agent arm looking ats aspirin, and the P2Y12 inhibitors clopidogrel, ticagrelor or prasugrel (45). PEAC aims to test the efficacy of aspirin in shortening clinical recovery time. The impact of aspirin on all-cause mortality among hospitalized patients is also under evaluation in the largest adaptive platform RCT for COVID-19 (RECOVERY) with 20,000 participants (46). RESIST (CTRI/2020/07/026791) aims to evaluate the role of aspirin plus atorvastatin in clinical deterioration characterized by progression to WHO clinical improvement ordinal score in 800 hospitalized patients with COVID-19 (47). CAM-Covid-19 evaluates the impact of higher dose of aspirin (325 mg QID) along with colchicine and montelukast on inflammatory markers such as hs-CRP in 34 patients. PARTISAN (NCT04445623) will be comparing the effect of prasugrel versus placebo among 128 patients with COVID-19 on the primary outcome of improved oxygenation expressed as the PaO2/FiO2 ratio at 7-day follow-up. Some RCTs are evaluating the impact of dipyridamole in hospitalized patients with COVID-19. Dipyridamole 100 mg QID and the combination of dipyridamole ER 200mg/ aspirin 25mg are being evaluated in 3 small RCTs (TOLD, DICER and ATTAC-19) for primary outcomes such as D-dimer level changes (for the first two trials) and improvement in COVID-19 WHO Ordinal Scale (a scale indicting severity of illness, from 0 [not infected] to 8 [death]) (ATTAC-19).

The use of DOACs in hospitalized ward patients with COVID-19 is under investigation in 5 RCTs. Low-intensity rivaroxaban is being investigated in 650 planned participants in the ACOVACT and XACT trials of hospitalized patients to assess outcomes such as all-cause mortality, ICU admission, and intubation. High-intensity (but not loading-intensity) DOACs including rivaroxaban and apixaban are being evaluated in large RCTs that will enroll a total of 4,750 participants (ACTION, COVID-PREVENT, FREEDOM COVID, and XACT).

C-19-ACS is an adaptive RCT conducted to evaluate the impact of the combination of low-dose rivaroxaban (2.5 mg BID) plus aspirin 75 mg/day plus clopidogrel 75 mg/day along with atorvastatin and omeprazole on 30-day all-cause mortality in 3,170 hospitalized patients with COVID-19. Patients with definite acute coronary syndromes are excluded from this RCT. The effect of dual pathway inhibition using the combination of low-dose rivaroxaban and aspirin is being evaluated in the adaptive ACTCOVID19 inpatient study. In this RCT of 4,000 patients, the rate of invasive mechanical ventilation or death is assessed at 45 days post-randomization.

Twenty-eight ongoing studies are being conducted to examine the efficacy of heparin-based regimens on primary outcomes such as all-cause mortality, venous and arterial thrombosis, re-hospitalization, the need for invasive mechanical ventilation, or composite outcomes inclusive of these in hospitalized patients with COVID-19. The majority of these RCTs has chosen standard-dose prophylactic anticoagulation regimen as the comparator. Intermediate-dose anticoagulation will be tested in DAWn-Antico (48), X-COVID-19, COVID-19 HD, COVI-DOSE, EMOS-COVID, NCT04360824, and EUCTR2020-001891-14-ES with 4,434 patients in total. On the other hand, a total of 18 RCTs with 19,776 patients will evaluate the efficacy of therapeutic anticoagulation in non-ICU hospitalized patients (49). Only two trials totaling 494 patients (IMPACT and HEP-COVID) will directly compare therapeutic and intermediate doses of heparin. The different intensities of heparin derivatives are summarized in Supplemental Table 3.

Recognizing that heparin has more than an anticoagulant effect but also an antiviral and anti-inflammatory effect, INHALE-HEP and NEBUHEPA are evaluating the impact of nebulized UFH on the rate of intubation in 856 hospitalized patients with COVID-19. PACTR202007606032743 evaluates the impact of nebulized UFH on PaO2/FiO2 ratio in 100 hospitalized patients. Standard of care (for INHALE-HEP and PACTR202007606032743) and standard-dose prophylaxis with LMWH (NEBUHEPA) are the comparators, respectively.

High mobility group box protein 1 (HMGB1) is a protein involved in the pathogenesis of inflammation. Elevated levels of HMGB1 is associated with worse the outcomes in COVID-19 (50). Dociparstat, a heparin derivative with presumed anticoagulant and anti-inflammatory properties, inhibits HMGB1 and may reduce the formation of NETs and the risk of thrombosis. The drug is being studied in NCT04389840 to assess its impacts on all-cause mortality and need for mechanical ventilation in 600 patients with severe COVID-19 (51).

Nafamostat is a synthetic serine protease inhibitor with anti-viral, anti-inflammatory, and anticoagulant activity previously used for anticoagulation during hemodialysis (52). Nafamostat is under evaluation in hospitalized patients with COVID-19 in 7 RCTs with 826 individuals in total. The primary efficacy outcome in most (5/7) of these trials is time to recovery.

The strengths of many of the antithrombotic trials among inpatients with COVID-19 include relatively large sample sizes and ample follow-up for detection of events. With multiple large clinical trials underway, robust evidence should soon be available comparing the intermediate/therapeutic doses of heparinoids versus usual care. However, studies such as PARTISAN and NCT04420299 have relatively small sample sizes and short periods of follow-up (7 days and 10 days, respectively), rendering them susceptible to type II error. There is also variability across the trials in methods for identification and ascertainment of thrombotic outcomes. Lack of blinding and blinded outcome adjudication are practical limitations for some of these trials.

### Ongoing Clinical Trials of Antithrombotic Agents in Critically-Ill Patients

The risk of thrombotic events appears to be highest among critically-ill patients with COVID-19. A systematic review estimated that VTE event rates in critically-ill patients with COVID-19 would be estimated at 27.9% (95% CI, 22.1%-34.1%) (6). Currently, there are 33 ongoing RCTs evaluating the role of antithrombotic agents in critically-ill patients with COVID-19 of which 18 RCTs enroll mixed non-ICU and ICU populations and 16 RCTs solely enroll ICU patients. The sample size of these studies range from 15 to 20,000 patients. These trials study the role of antiplatelet agents (aspirin, clopidogrel and dipyridamole), systemic anticoagulants (intermediate to full-therapeutic-dose of heparin and DTIs), inhaled UFH, fibrinolytic agents (tenecteplase and alteplase), and nafamostat. Inclusion criteria in 11 of 34 RCTs require D-dimer cut-offs ranging from >500 ng/ml to >3,000 ng/ml (or defined as >2-6 times the upper limit of normal limit). All-cause mortality, venous and arterial thrombotic complications, and oxygenation (expressed mostly as PaO2/FiO2) status are the most common components of the primary efficacy outcomes. Bleeding complications are the most widely used primary safety outcome among these studies.

The role of antiplatelet agents is under investigation in critically-ill patients in 4 trials. As previously described, dipyridamole (TOLD) and aspirin (RECOVERY and CAM-Covid-19) are under evaluation. COVID-PACT is a multicenter, open-label study that will randomize 750 patients with 2 x 2 factorial design trial to full-dose anticoagulation vs. standard-dose prophylactic anticoagulation with heparin-based regimens (first randomization), and to antiplatelet therapy with clopidogrel vs. no antiplatelet therapy (second randomization). The primary efficacy outcome is the incidence of VTE or arterial thrombosis incidence 28 days after enrollment.

UFH and/or LMWH (19 studies) are the most common antithrombotic regimens under investigation in the ongoing trials in critically-ill patients. INSPIRATION, IMPROVE, DAWn-Antico, and COVI-DOSE are testing intermediate-dose versus standard prophylactic dose anticoagulation in more than 1,500 participants in total. INSPIRATION has recently completed enrollment of 600 patients and the results are imminent (53). Eleven RCTs are evaluating the potential role of therapeutic-dose versus standard prophylactic dose anticoagulation in 5,142 patients. In December 2020, preliminary results of an interim analysis of pooled critically ill patients enrolled in the three trials (ACTIV-4a, REMAP CAP, and ATTACC) by the Data Safety and Monitoring boards paused enrollment of critically ill patients due to futility for the endpoint of freedom from organ support at 21 days. Enrollment of moderately ill patients in this composite trial continues. This decision was based on the determination of futility, and a potential for harm due to possibly higher rates of bleeding. More details are forthcoming (40). Finally, IMPACT and HEP-COVID are comparing therapeutic anticoagulation with intermediate-dose anticoagulation in a total of 494 individuals.

The only published RCT in critically-ill patients with COVID-19 is HESACOVID, a single-center study of 20 patients requiring invasive mechanical ventilation randomized to therapeutic versus standard-dose anticoagulation. Therapeutic-dose anticoagulation significantly increased PaO2/FiO2 and ventilator free days (15 days [IQR 6–16] versus 0 days [IQR 0–11], p = 0.028) (38). The study did not have sufficient power to compare all-cause mortality between the study groups. Bleeding may have been underestimated due to barriers of performing imaging testing, including computed tomography to identify a source, in critically-ill patients (54).

CHARTER-Irl, CHARTER-MT, and NCT04397510 are evaluating the utility of nebulized UFH in 292 mechanically-ventilated critically-ill patients with COVID-19. The primary outcome for CHARTER-Irl is the alterations in D-dimer area under the curve (AUC) within a 10-day follow-up, and for CHARTER-MT is ventilator-free days with a follow-up duration of 28 days; the primary outcome for NCT04397510 is improvement in PaO2/FiO2 ratio within 10 days.

The use of parenteral anticoagulants other than UFH and LMWHs in COVID-19 is being studied in two trials. IMPACT will randomize 100 ICU patients with COVID-19 into 4 arms to compare fondaparinux, argatroban, intermediate-dose heparin and therapeutic-dose heparin (UFH/LMWH) with the primary outcomes of 30-day mortality. In ANTI-CO, bivalirudin is being investigated in 100 critically-ill patients for the primary outcome of improvement in oxygenation as determined by the PaO2/FiO2 ratio (55).

There are six RCTs (AtTAC, STARS, TRISTARDS, TACOVID, NCT04505592, and IRCT20200415047080N1) evaluating the safety and efficacy of fibrinolytic therapy (tenecteplase or alteplase) on COVID-19 related respiratory failure in a total of 440 patients (56). Most of these trials include patients with severe disease (severe acute respiratory distress syndrome, elevated troponin levels, and elevated D-dimer levels). The primary outcomes in 5 of these trials include the improvement in PaO2/FiO2 ratio or ventilator free days. The time frame for studies evaluating the change in PaO2/FiO2 ratio is between 48-72 hours and for those which evaluate ventilator free days is 28 days. Patients receiving therapeutic anticoagulation, and those with thrombocytopenia, or history of intracranial or gastrointestinal bleeding are excluded from fibrinolytic therapy trials.

Nafamostat is under evaluation in 4 studies in critically-ill patients with COVID-19 (DEFINE, RACONA, NCT04418128, and NCT04623021) with 650 participants in total.

Research in the ICU faces several challenges for study design, data/sample collection, and patient follow-up (57). In many cases, patients are unconscious and obtaining informed consent requires discussion with healthcare proxies. This is further complicated because visitors are prohibited. The strengths of the aforementioned studies in the ICU include the diversity of studied antithrombotic agents, and sample size in many RCTs. There are also a number of notable limitations to these trials. The most important limitation is the small sample size in several studies, raising the possibility of type II error. The small sample size will mostly influence trials of thrombolytic therapy and non-heparin anticoagulants.

### Ongoing Clinical Trials in Post-Discharge Patients

ACTIV-4c (NCT04650087) is a double-blind, placebo-controlled RCT which will evaluate the impact of apixaban 2.5 mg BID on the rate of all-cause mortality, and arterial and venous thromboembolism on 5,320 post-discharge patients. MICHELLE (NCT04662684) is an open-label RCT with 320 participants aims to evaluate the safety and efficacy of rivaroxaban 10 mg QD for 35+/-4 days versus no intervention after hospital discharge with a composite efficacy outcome of VTE and VTE-related death.

In addition, there are 7 RCTs with a projected total of 1,452 participants which will continue the already assigned antithrombotic therapy after discharge in patients who were randomized in the general medical wards or in the ICU. In COVID-PREVENT, XACT, and NCT04508439 RCTs, post-discharge thromboprophylaxis with rivaroxaban (10 or 20 mg QD) is being investigated in 680 participants who were enrolled in general medical wards to measure the incidence of VTE at 30-35 days after discharge. In the HERO-19 study, edoxaban 60 mg QD or placebo will continue after discharge in 172 patients who were randomized in the ICU or non-ICU settings to evaluate all-cause mortality rate and VTE incidence at 42 days. In the INSPIRATION study, intermediate or standard prophylactic dose of enoxaparin will be continued after discharge in 600 patients who were randomized in the ICU to evaluate the rate of VTE. Finally, aspirin in the PEAC study, and dipyridamole ER plus aspirin in the ATTAC-19 study, will be continued after discharge in patients randomized in the non-ICU general wards.

### Incorporation of Vulnerable Populations in the Ongoing Trials

Most of the ongoing RCTs are excluding patients at increased risk of bleeding, or with acute and chronic hepatic failure. In more than 50% of the trials designed to evaluate escalated dose anticoagulation, patients with CrCl < 15 mL/min are excluded. CoV-Hep is an open-label study which evaluates the role of low-dose (10 IU/kg/h) intravenous UFH on the rate of clotted dialyzers in 90 critically-ill patients with COVID-19 undergoing continuous venous-venous hemodialysis with a follow-up duration of 3 days. Specific dose adjustment for obesity is considered for 10 out of 34 trials of systemic heparin compounds. Pregnant women are excluded from 25 out of 34 trials of systemic heparin compounds. While patient selection in these studies is based on practical considerations, it is unlikely that high-quality evidence will soon be available for antithrombotic therapy in such vulnerable subgroups (Figure 4 and Supplemental Figure 2).

**Figure 4.**
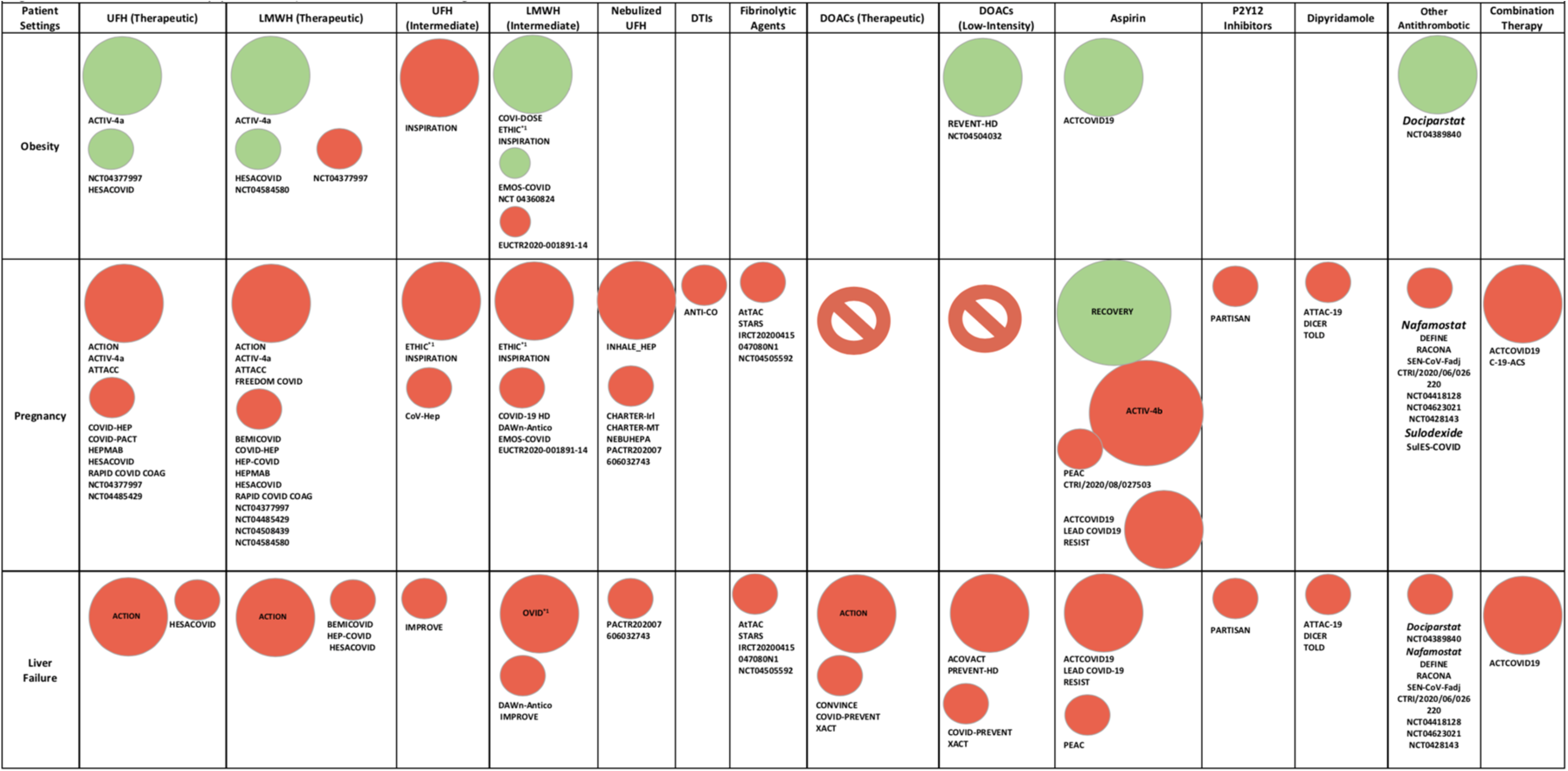
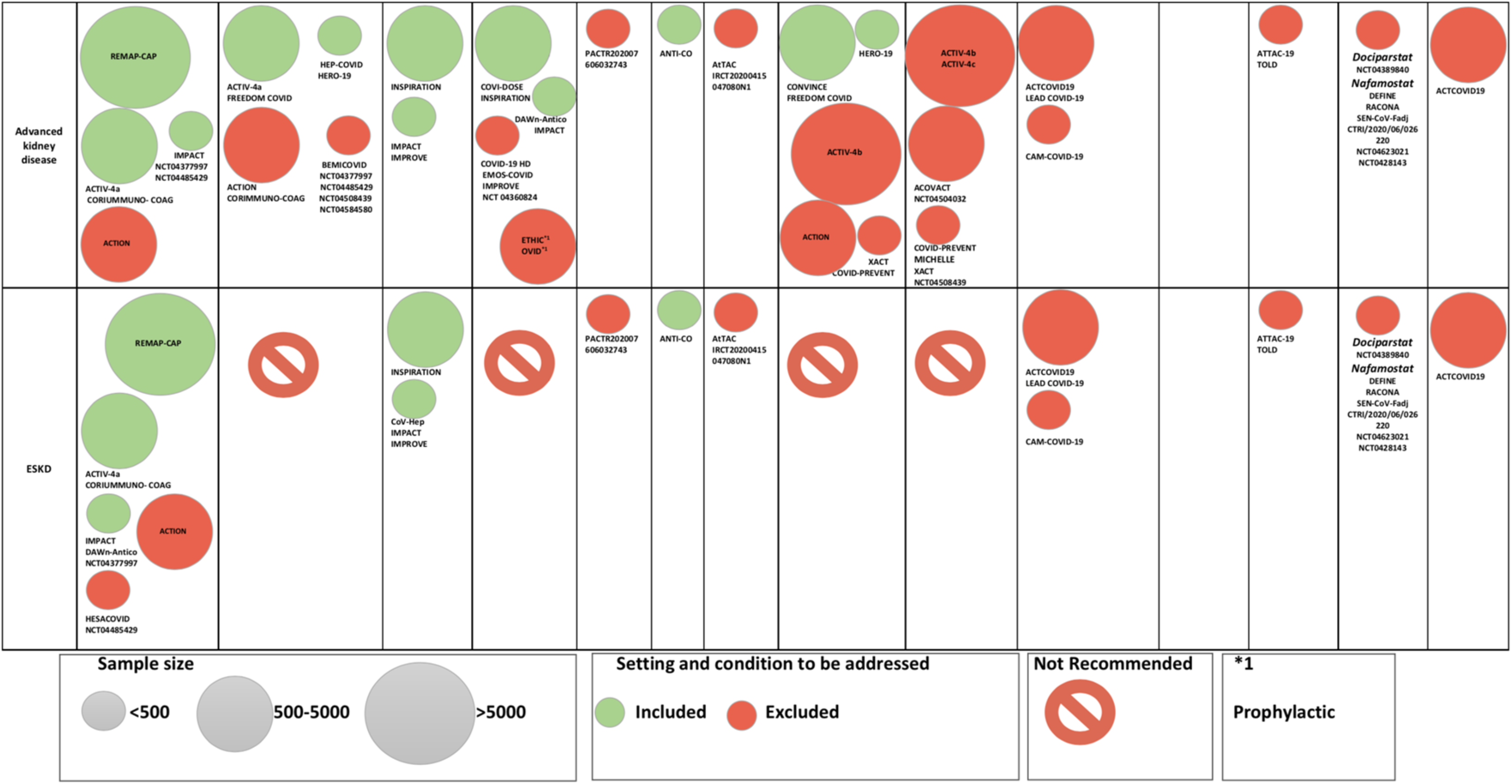
Illustration of how vulnerable populations were/ were not included in the existing trials. Caption: Categorizing the RCTs evaluating different agents in vulnerable populations including patients with advanced kidney disease, ESKD, patients with liver failure, and obese patients. Further details are illustrated in Supplemental Figure 1. Obesity is defined differently in different RCTs; body mass index greater than 30, 35, and 40 kg/m2 and weight more than 100 and 120 kg are among the most used definitions among RCTs. Others: dociparstat, nafamostat, and sulodexide. DOAC: Direct Oral Anticoagulants, DTI: Direct Thrombin Inhibitors, ESKD: End Stage Kidney Disease; LMWH: Low Molecular Weight Heparin; RCT: Randomized Clinical Trials; UFH: Unfractionated Heparin.

### The Impact of RCTs on the Future Practice of Antithrombotic Therapy

A large number of RCTs will help to delineate the efficacy and safety of antithrombotic agents in patients with COVID-19 (Central Illustration). Until the results accrue, participation in these RCTs is encouraged. Efficacy outcomes vary based on the location of enrollment; i.e., between outpatient trials and inpatient trials. As for safety outcomes, many of the trials are systematically assessing major bleeding using the International Society on Thrombosis and Haemostasis (ISTH) criteria or the Bleeding Academic Research Consortium definitions (58, 59). While observational evidence suggests low rates of major bleeding, (33, 60), observational studies have the potential for underreported outcomes and therefore RCTs with systematic and prospective capture of both thrombotic and bleeding events will help determine the true risk-benefit ratio for treatments. This is especially the case since risk factors for thrombosis in COVID-19, such as D-dimer, may also predict bleeding (33).

While results from the individual trials may inform interim practice, some challenges persist. The large number of antithrombotic agents under investigation, the variable dosing regimens tested, and variability in trial conduct as well as methods of outcome detection and adjudication may complicate the identification of the optimal regimens. A prospective meta-analysis of RCTs, ideally with individual participant data, will help to assess the effects of distinct agents across the spectrum of disease severity and may address the clinical and statistical heterogeneity of the upcoming results. Efforts to harmonize endpoints have been advocated, with creation of common data elements for VTE for example, to aid in pooling trial results (61, 62). In addition, there are few head-to-head comparisons for many of the experimental therapies, such as intermediate-dose compared with fully-therapeutic heparin-based regimens. Network meta-analytic techniques might generate insights into the comparative tradeoffs of these regimens (63). Additional biomarker and clinical risk prediction sub-studies can also further elucidate subgroups with more favorable net benefit profiles from distinct regimens. Moreover, the remaining knowledge gaps summarized in Table 1 should be kept in mind so that the design of additional studies could be considered.

**Table 1.** Expected Knowledge Gains from Ongoing Antithrombotic Therapy Trials in COVID-19 and Ongoing Knowledge Gaps.

|  |  |  |  |  |
| --- | --- | --- | --- | --- |
|  | Outpatient | Non-Critical Inpatient | ICU | Post-Discharge |
| Expected Knowledge Gain | - The safety/efficacy of **standard prophylactic doses** of LMWHs and DOACs compared to standard of care or placebo in high risk patients with early stage of the COVID-19<br>- The impact of **aspirin** administration on rate of MACE, disease progression, hospitalization, and death in patients with acute, symptomatic COVID-19<br>- The safety/efficacy of **sulodexide** in patients with acute, symptomatic COVID-19 | -The safety/efficacy of **intermediate dose** and **therapeutic dose heparin derivatives** or DOACs compared to standard prophylactic anticoagulation<br>- Proof of concept data of the role of **inhaled antithrombotic therapy** in patients with COVID-19<br>- The impacts of **antiplatelet agents** on all-cause mortality<br>- The safety/efficacy of **dociparstat** and **nafamostat** in hospitalized patients with COVID-19 | -The safety/efficacy of **intermediate dose** and **therapeutic dose anticoagulation** compared with prophylactic anticoagulation<br>-Effects of short-term infusion of **bivalirudin** on PaO<sub>2</sub>/FiO<sub>2</sub> ratio<br>- The impacts of **antiplatelet agents** on thrombotic outcomes and mortality<br>- Proof of concept data on the role of **fibrinolytic therapy** in critically ill patients<br>- Proof of concept data of the role **inhaled antithrombotic therapy** in mechanically ventilated patients with COVID-19 | -The safety/efficacy of extended anticoagulation with **DOACs** or **LMWHs** after hospital discharge |
| Remaining Knowledge Gap | - PMA needed to understand the relative efficacy of **antiplatelet agents, standard prophylactic dose of enoxaparin** in comparison with DOACs | - The efficacy/safety of **fondaparinux, DTIs, and danaparoid** compared to standard prophylactic anticoagulation<br>- PMA needed to understand the tradeoffs of various investigational antithrombotic regimens<br>- **Best assay for LMWH/UFH monitoring** in non-critically ill patients with COVID-19 | - The impact of **antiplatelet therapy** on survival in critically ill patients with COVID-19<br>- PMA needed to understand the tradeoffs of various investigational antithrombotic regimens<br>- **Best assay for LMWH/UFH monitoring** in critically ill patients with COVID-19 | -The role of **antiplatelet agents** on VTE incidence in post-discharge patients |
| COVID-19: Coronavirus Disease 2019, DOAC: Direct Oral Anticoagulants, DTI: Direct Thrombin Inhibitors, ICU: Intensive Care Unit, MACE: Major Adverse Cardiovascular Events, PMA: Prospective Meta-Analysis, UFH: Unfractionated Heparin, VTE: Venous Thromboembolism. |  |  |  |  |

Anti-inflammatory properties and activity against thromboinflammation have been attributed to several antithrombotic regimens including heparin derivatives and antiplatelet agents (30, 64, 65), with the potential to reduce large-vessel thrombosis and improve outcomes. Another evolving concept is the role of microthrombosis and pulmonary intravascular coagulopathy (7, 8, 66) in the pathophysiology of respiratory failure in COVID-19 (67). Results from the small HESACOVID study suggested improved arterial oxygenation (PaO2/FiO2) with therapeutic versus standard-dose prophylaxis anticoagulation in critically-ill patients with COVID-19 (38). However, combined investigation of three large-scale randomized trials of therapeutic anticoagulation (ACTIV-4a, REMAP CAP, and ATTACC) paused enrollment of critically ill patients for futility; we await further clarifications (40).

Therapeutic drug monitoring of the investigational agents is also important. Even when an agent is selected (e.g., UFH), the best method for dose titration or adjustment remains uncertain (68). Some experts recommend measuring anti-Xa levels in those receiving intravenous unfractionated heparin, since the high levels of Factor VIII observed among critically-ill patients with COVID-19 may interfere with aPTT assays. The necessity and optimal method for dosing and monitoring of heparins and LMWHs, in particular for patients with kidney disease or obesity is yet to be elucidated, and is even understudied outside COVID-19 (69). Ideally, future strategy trials should test the merits and limitations of these monitoring tests.

### Clinical Trial Enterprise During COVID-19 Pandemic: Is a Quantum Leap Taking Place?

The clinical trial enterprise has been significantly affected during the COVID-19 pandemic (70). Patient recruitment in many ongoing pre-COVID-19 trials was temporarily halted. Notable challenges such as barriers to follow-up and site-monitoring persist. However, the desire to provide an evidence-based response has been one of the key drivers of positive changes during the pandemic (71). These changes include multi-specialty study teams, harmonization of multicenter protocols, expedited multi-institutional agreement execution and institutional review board and governmental agency approvals, accelerated informed consent, and enrollment with digital contact-free technology, expeditious outcomes ascertainment, remote monitoring, and dissemination of the findings via fast-track publications, pre-prints, and social media accounts from scientific societies or investigators (Table 2) (72–74).

**Table 2.** Antithrombotic Therapy Trial Design Before and During the COVID-19 Pandemic.

|  |  |  |
| --- | --- | --- |
| Various Areas of Clinical Research | Before COVID-19 Pandemic | During COVID-19 Pandemic |
| **Investigators** | • Single specialty-based collaboration common<br>• Focused, often established study groups | • Specialty-based and multi-specialty collaboration common<br>• Frequent ad hoc collaborations within and between institutions and countries |
| **Study Design** | • Diverse research priorities<br>• Patient enrollment over a long time period, recruitment time could be slow of fast<br>• Long-term follow up a routine feature of many trials | • Distinct focus on COVID-19-related trials, some adaptations required for pre-COVID-19 trials.<br>• Time-sensitive trial design (to provide rapid access to high quality evidence). Trial design in short period of time may lead to multiple smaller and underpowered trials rather than larger multicenter collaborations<br>• Urgently needed medical solutions necessitate relatively fast patient enrollment<br>• Incorporation of pragmatic design features.<br>• Multiple projects around the world occasionally leading into several smaller trials rather than fewer large-scaler trials<br>• Short-term follow up most common<br>• Applicability for adaptive platform design for multiple aspects of COVID-19 trials. Multiple interventions, quick enrollment, and the possibility of re-estimation of the optimal sample size during the study<br>• Higher certain event rates (death or re-admission) that excepted from protocol |
| **Funding/ Financial Support** | • Time-consuming review and approval process for funding allocation | • Accelerated review, prioritizing trials that impact the response to the pandemic |
| **IRB Approval** | • Time-consuming process with occasional long delays prior to approval | • IRBs meeting more frequently, often resulting in rapid review and approval<br>• More permissive regulations may expedite trial initiation |
| **Informed Consent** | • Based on paper forms, may be cumbersome | • In-person or remote electronic informed consent available in many trials |
| **Participants Enrollment and Engagement** | • Variable willingness for trial participation by patients | • Patients willing to participate and engage in trials as partners<br>• Periodic slowdown or interruptions in enrollment for some non-COVID-19 trials, in COVID-19 trials there may be changes in enrollment rate with COVID-19 disease waves. |
| **Monitoring and Auditing** | • On site session for multiple predefined monitoring visits<br>• On site or in-person data audits | • Frequent off-site online sessions with more restricted onsite visit<br>• Remote monitoring and follow up<br>• Rapid enrollment makes keeping up to date with monitoring difficult, with risk of greater numbers of protocol deviations going un-noticed.<br>• Ascertainment of events other than all-cause mortality may be challenging due to limitations in testing strategies during the pandemic |
| **Clinical Events Adjudication** | • Central blinded outcome adjudication common<br>• Face to face meetings<br>• High costs<br>• Time consuming process to request ad-hoc data from sites, summarize, and send back to adjudication meetings | • Some trials not able to incorporate endpoint adjudication (not recommended if resources allow)<br>• Systematic and blinded adjudication in online meeting for assessment endpoints<br>• Remote periodic meetings<br>• Less expensive and quicker than face to face adjudication |
| **DSMB Meetings** | • Face to face meetings<br>• High costs | • Many trials using online platforms for DSMB meetings<br>• Less costly |
| **Follow-Up** | • Face to face visits or phone calls<br>• More costly | • Remote monitoring and follow-up in many trials by phone calls and use of digital technology<br>• Cost-saving and more efficient |
| **Dissemination of Results** | • Longer peer-review process<br>• More strict criteria for publication<br>• Uncommon use of pre-print servers | • Fast-track peer-review process expedites the dissemination of completed studies. However, very quick peer-review has occasionally missed important flaws of submitted reports.<br>• Frequent use of pre-print servers to share the early results of the studies. The benefits of rapid dissemination and potential limitations with lack of peer-review should be considered among the audience of the results. |
| APT: Adaptive platform trials, COVID-19: Coronavirus Disease 2019, DSMB: Data and Safety Monitoring Board, IRB: Institutional Review Board, RCT: Randomized Clinical Trials, RFA: Request for Application. |  |  |

Although traditional RCTs have provided a great deal of knowledge for modern medicine, they are confined to testing a limited number of interventions. Since COVID-19 has multi-organ involvement and broad manifestations (including inflammation, ARDS, thrombosis and others), adaptive platform trials –which allow for testing multiple interventions in a single disease based on a decisive algorithm– have gained attention (75). This type of trial has a perpetual and multi-arm-multi-stage design.(76) The RECOVERY Trial (77) and the World Health Organization Solidarity Trial (78) have tested different steroid and antiviral regimens, respectively, and have some additional agents under investigation, including aspirin in one of the hypotheses from RECOVERY. REMAP-CAP is testing several interventions including steroids, antivirals, biologic agents, simvastatin, and antiplatelet therapy. ACTIV4 platform is similarly using an adaptive design for antithrombotic agents.

Notwithstanding the good will of investigators, the constant pressure to provide rapid pandemic response may pose challenges, as well. In some cases, multiple small single-center RCTs underpowered for their clinical points or using surrogate endpoints with short follow-up have been designed (71, 79) and may compete against larger multicenter, and potentially more definitive, studies. The large numbers of these trials alone, in addition to the intense pressure to present broadly and publish these findings suggests at least some potential for Type I error with amplification of these results through rapid dissemination of the results.

Additional methodological aspects deserve attention. Interpretation of these trial results may be limited by underutilization of placebo (perhaps except for the outcome of mortality) (54, 79). Some experts consider that the pressures of working during a global pandemic makes the use of placebo more aspirational than realistic. Nevertheless, when feasible, placebo control improves the internal validity of a trial. Further, appropriate endpoint assessment, including blinded adjudication when feasible, and pre-specified analysis methods will remain of importance (54).

IRBs and independent ethics committees may experience the burden of numerous protocol submissions and amendments during the pandemic. Burnout of healthcare systems during the pandemic, and the risks to the research teams are unique challenges that should also be considered when designing and executing study protocols (72). Investigators should attempt to foresee some of the challenges in order to minimize the need for protocol amendments (80–82). Moreover, the informed consent process has become adapted to facilitate discussions by telephone or video conference, followed by verbal confirmation, and documentation of consent using approved software programs and electronic signature, where acceptable (80, 83).

Efficacy and safety outcomes monitoring is also critical. Execution of online Clinical Event Committee (CEC) and Data and Safety Monitoring Board (DSMB) meetings for assessing the adverse events is a fast safe, efficient alternative to face-to-face meetings. If done with appropriate planning to adhere to standards of high-quality CEC and DSMB meetings, such approaches may be considered even when society transitions out of the pandemic (81, 83).

Peer-review and dissemination of the studies has had unique challenges and advancements, too. Journal editors and reviewers have been pressured for rapid release of the results of completed studies. This has activated fast-track peer-review process more than ever. Despite its merits, the “COVID-19 fatigue” created by the fast-track review process might negatively impact the quality of peer-review, as noted by occurrence of post-publication major revisions and retractions, including in major journals (84). In a recent study, only 29% of the clinical trials of patients with COVID-19 reviewed on ClinicalTrials.gov met the Oxford Centre for Evidence-Based Medicine (OCEBM) level 2 evidence (85). The process of peer-review remains an imperfect –yet essential– step in the evaluation and reporting of results (86). Preprint servers include full drafts of research studies shared publicly before peer-review. Pre-prints have the potential benefit of early dissemination and opportunity for feedback and discussion, and could be of substantial benefit during the pandemic. With a preprint, key researchers in the field can discover findings sooner; indicate critical errors, or suggest new studies or data that strengthen the argument (87). The limitations of pre-prints should be also communicated transparently, so that similar weight is not placed on pre-print and peer-reviewed literature by the lay people, the press, healthcare workers, or policy-makers. Indeed many retracted papers were from pre-print servers (84).

Prospectively planned meta-analyses would be of particular help during the pandemic. Such studies can help understand the heterogeneity of the findings between interventions, between distinct studies, and within subgroups. Prospective meta-analysis can also help with pooled comparisons for interventions with small individual studies, as well as indirect comparisons for interventions that do not have sufficiently-large head-to-head comparisons in existing studies.

## CONCLUSIONS

Optimal antithrombotic therapy in patients with COVID-19 is yet to be determined. Results of these ongoing RCTs, and prospective meta-analyses of the completed studies will help clarify whether any of the plentiful antithrombotic regimens under investigation can safely mitigate thrombotic complications and improve patient outcomes.

## Data Availability

N/A

## ABBREVIATIONS AND ACRONYMS

COVID-19: coronavirus disease 2019
DOAC: direct oral anticoagulant
DTI: direct thrombin inhibitor
ICU: intensive care unit
LMWH: low molecular weight heparin
RCT: randomized controlled trial
SARS-CoV-2: severe acute respiratory syndrome coronavirus 2
UFH: unfractionated heparin
VTE: venous thromboembolism

## ACKNOWLEDGEMENTS

The authors would like to express their sincere gratitude to Fatemeh Esmaeili, MS, for her kind assistance in graphic designs.

## HIGHLIGHTS

- Venous and arterial thrombosis are among the prevalent manifestations of COVID-19.
- The optimal thromboprophylactic regimens still remain unknown in patients with COVID-19.
- At this time, dozens of RCTs are evaluating the role of antithrombotic regimens among outpatients and inpatients with COVID-19.
- COVID-19 has led to changes in the design, conduct, analysis, and reporting of the RCT results.

---

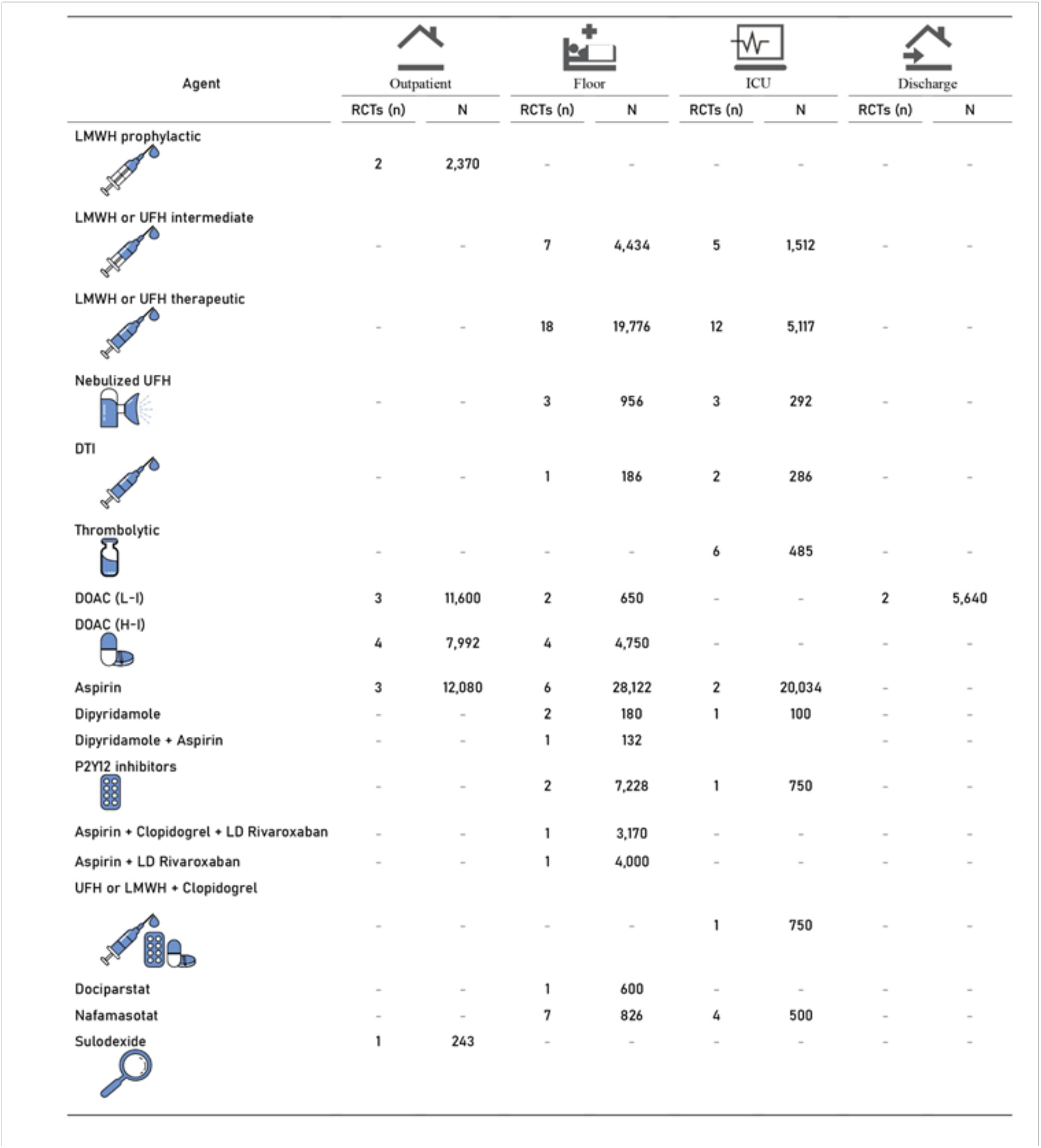
Central Illustration. Succinct Summary of ongoing RCTs of antithrombotic therapy in COVID-19. DOAC: Direct Oral Anticoagulant; DTI: Direct Thrombin Inhibitors; H-I: High-Intensity; ICU: Intensive Care Unit; L-I: Low-Intensity; LD: Low Dose; LMWH: Low Molecular Weight Heparin; N: Number of Participants; RCTs: Randomized Controlled Trials; UFH: Unfractionated Heparin.

**Supplemental Figure 1.**
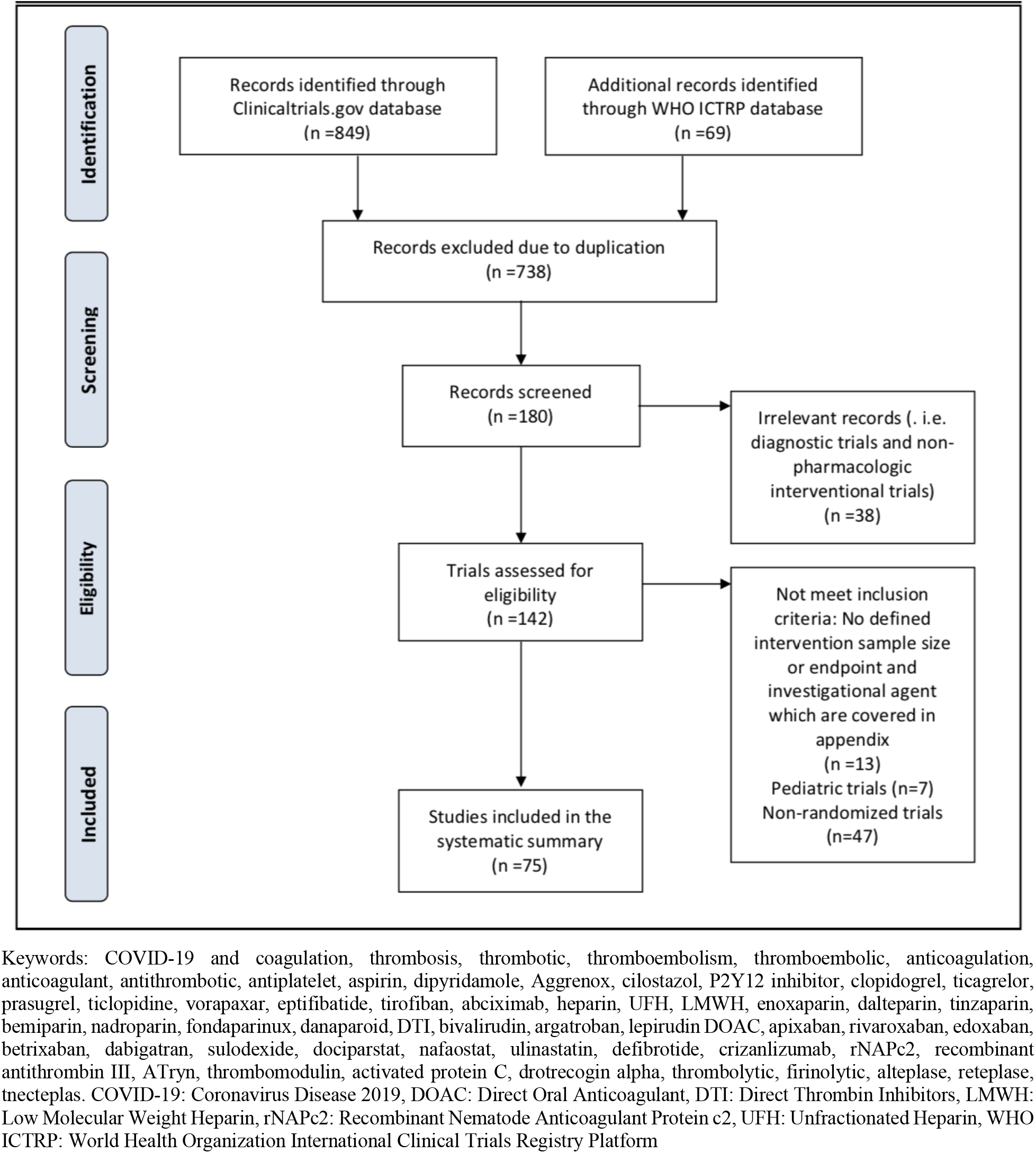
PRISMA Flow Diagram.

**Supplemental Figure 2.**
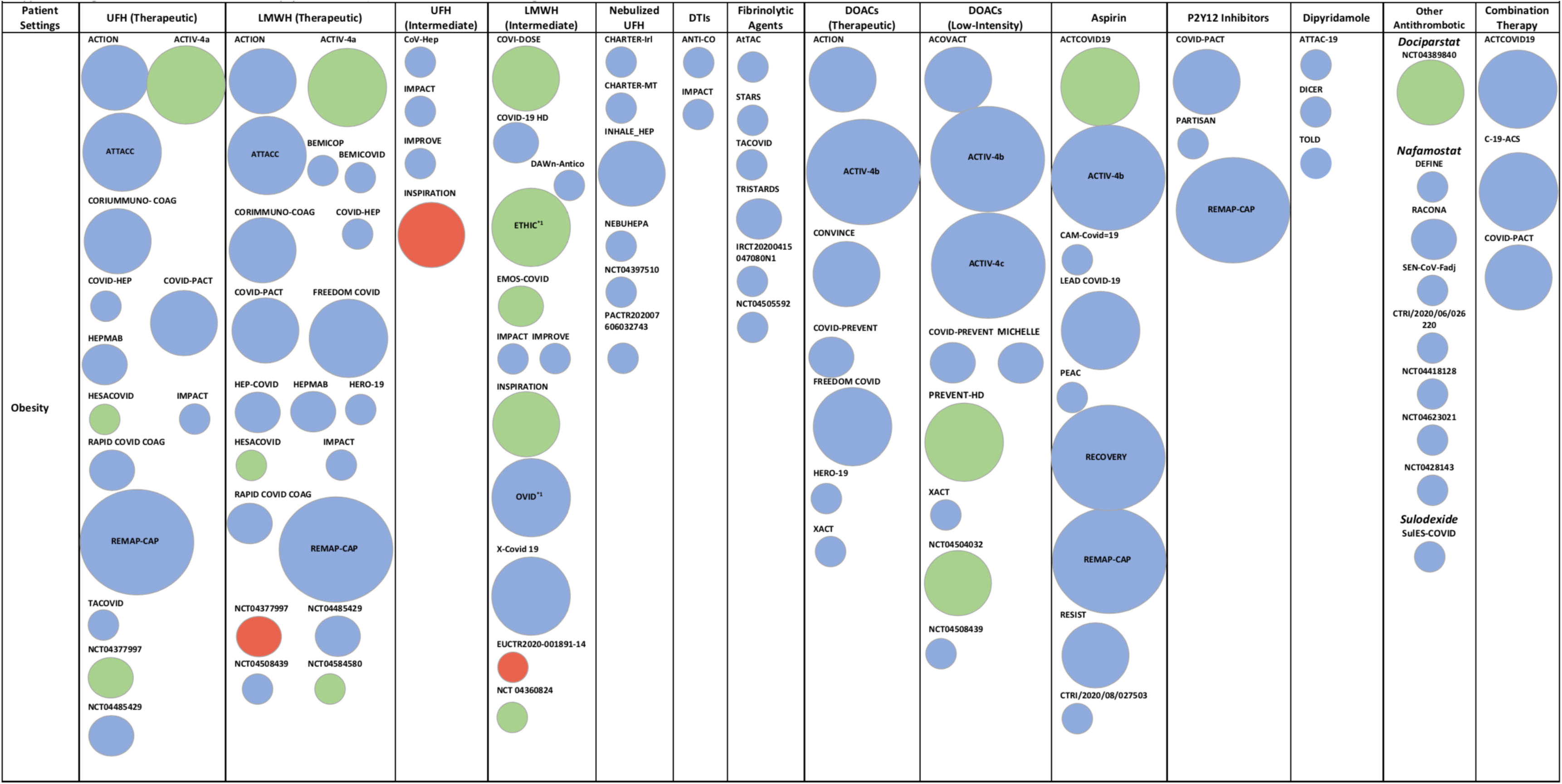
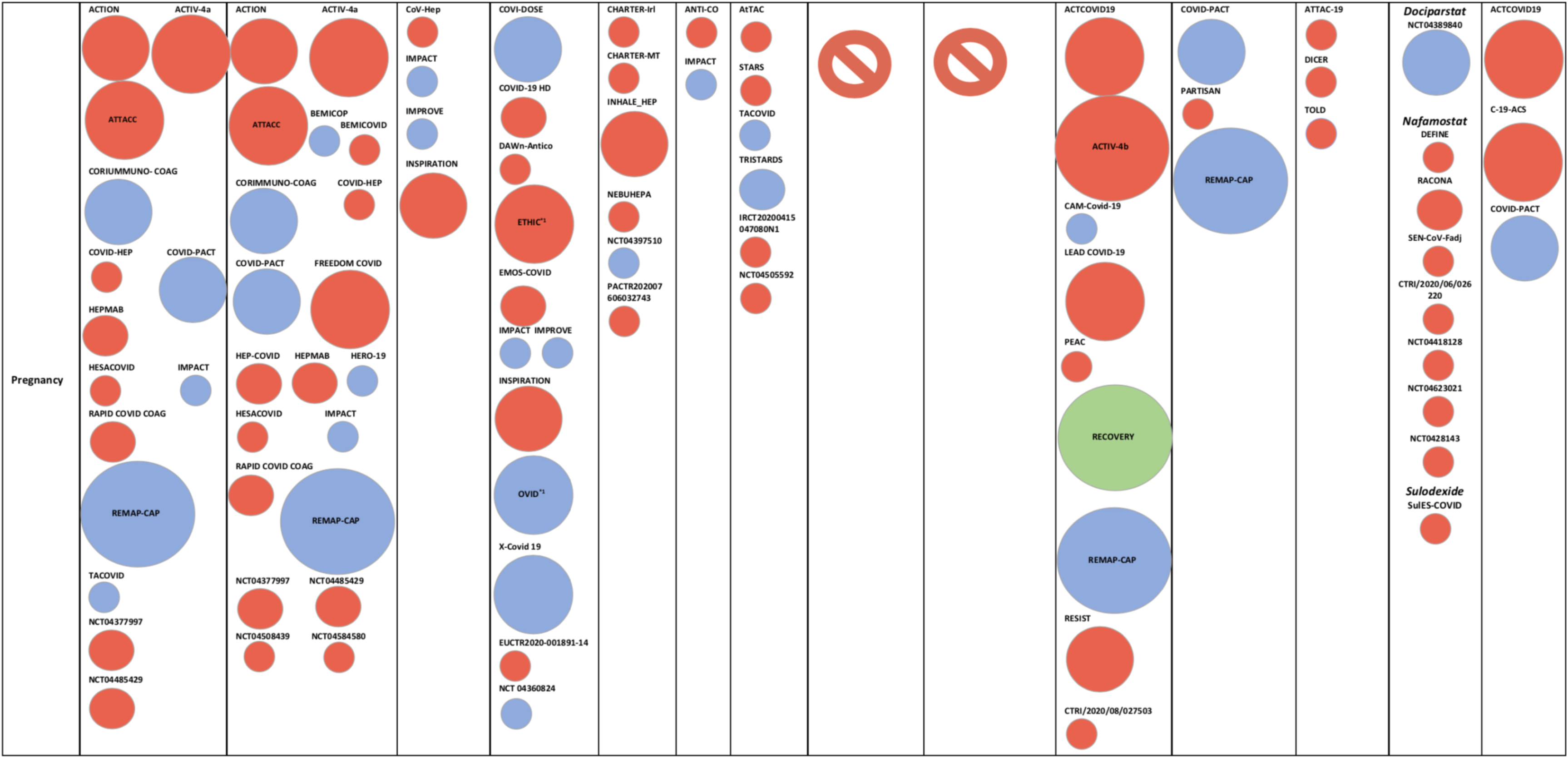
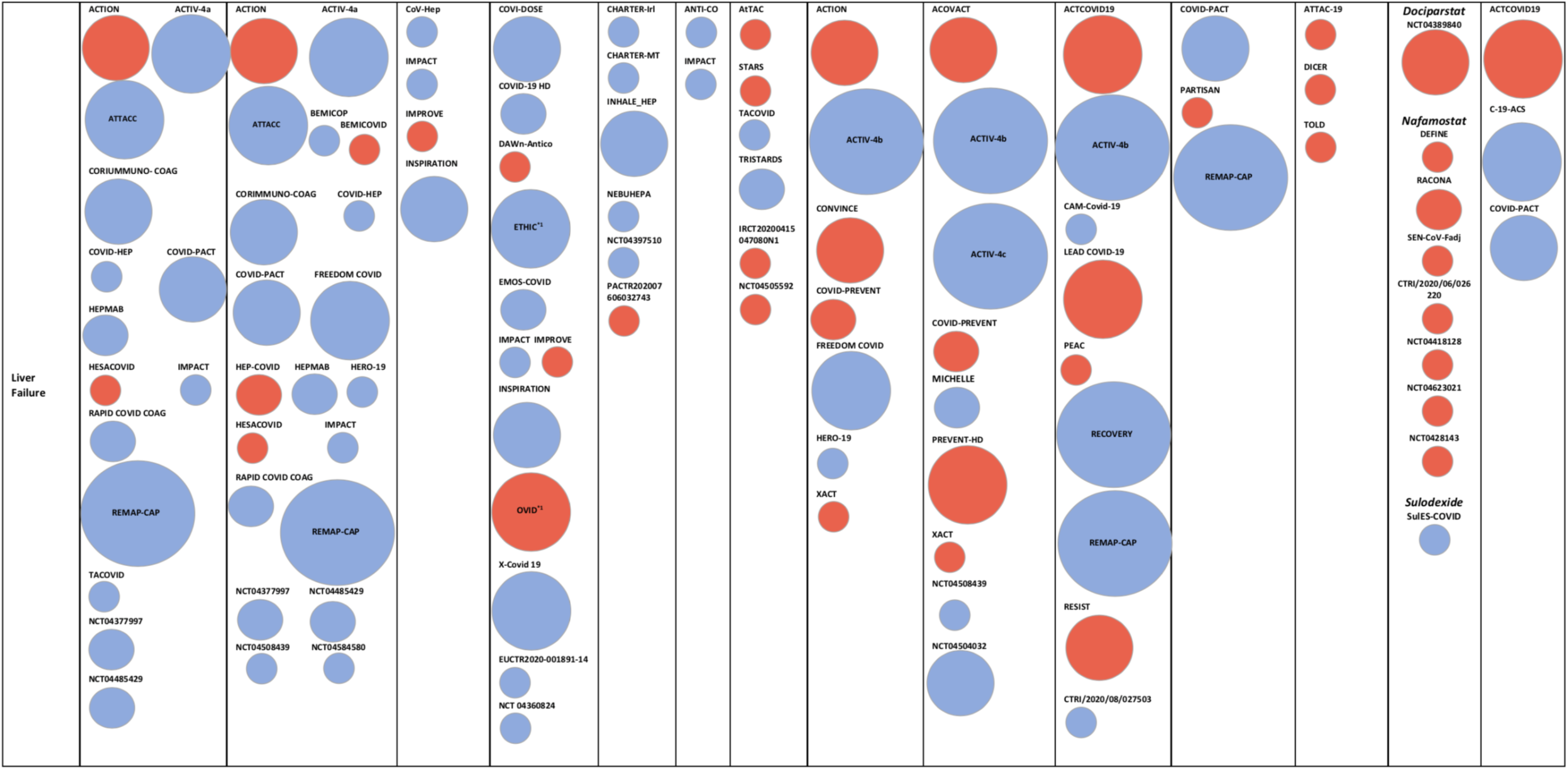
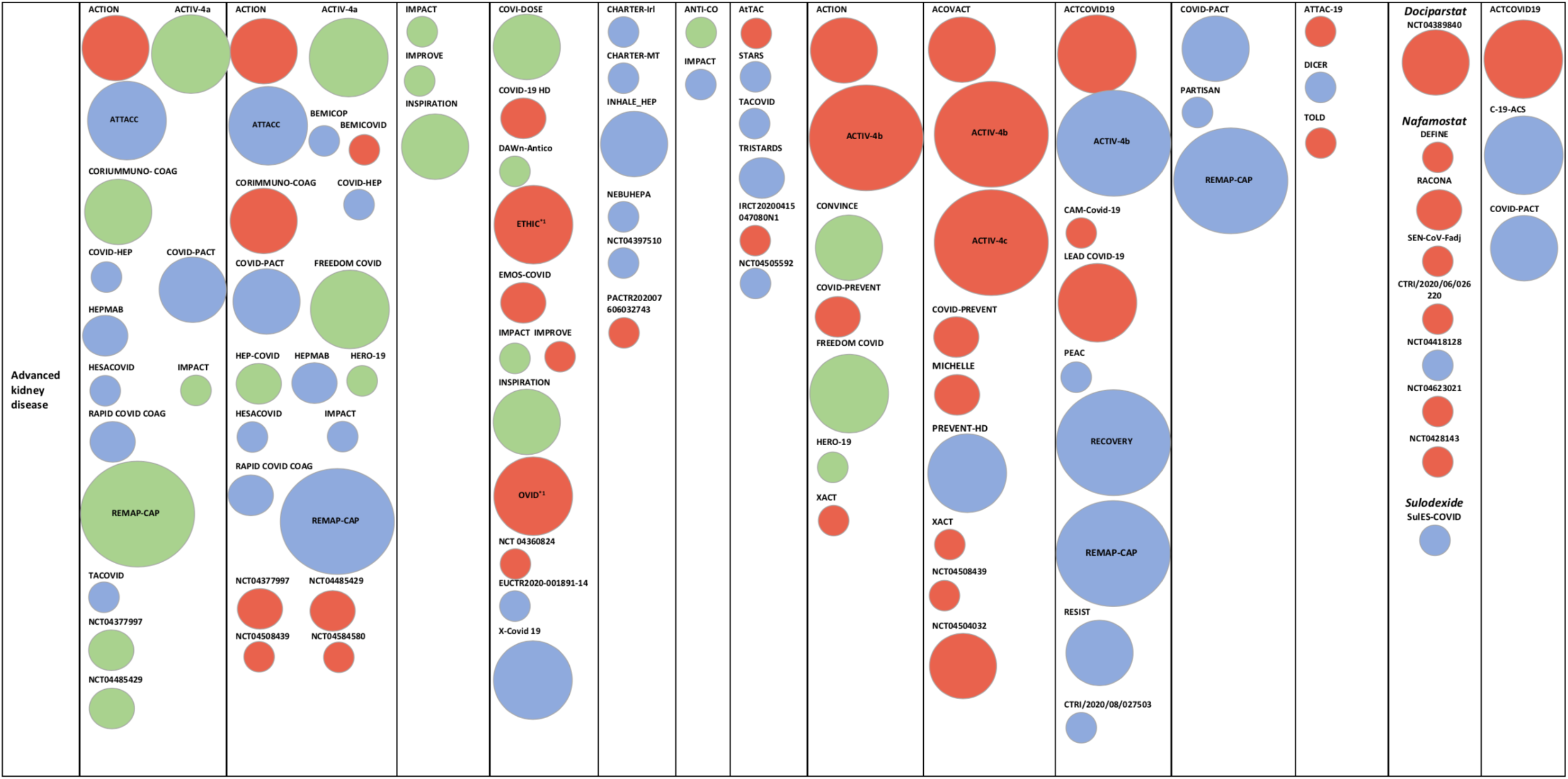
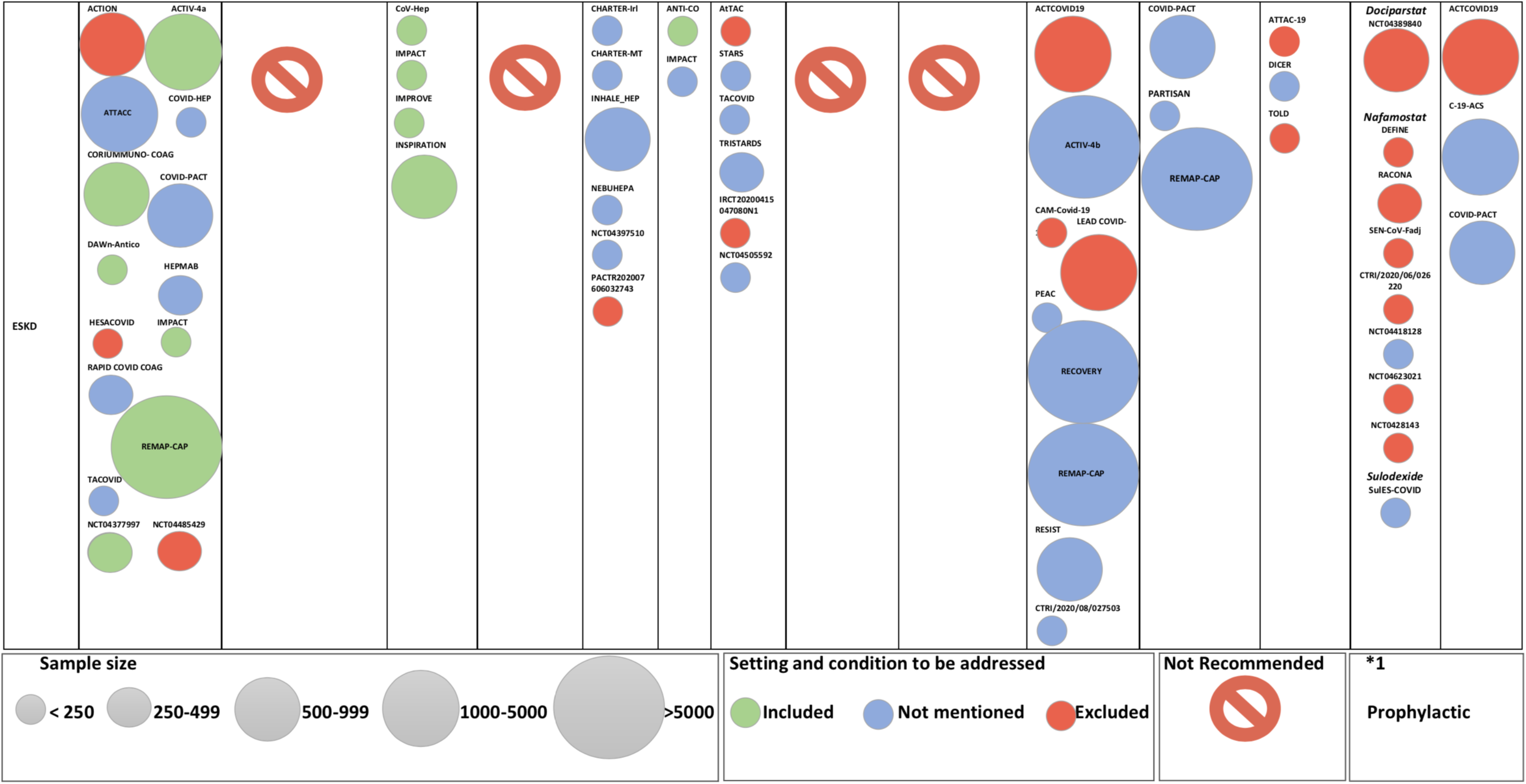
Illustration of how vulnerable populations were/were not included in the existing trials.

**Supplemental Table 1.** Study-level Inclusion and Exclusion Criteria for the Systematic Review.

|  |  |
| --- | --- |
| Inclusion Criteria | Exclusion Criteria |
| • Focused on patients with COVID-19 | • Studies of patients without COVID-19 |
| • Designed as a randomized controlled trial | • Non-randomized design |
| • Registered at clinicaltrials.gov or the WHO ICTRP (or identified via hand-searching by authors), date of last search: December 16, 2020 | • Minimum detail including the intervention, eligibility criteria and estimated sample size not available |
| • At least one active randomized arm including widely available antithrombotic agents such as antiplatelet agents, anticoagulants, fibrinolytic therapy (or investigational antithrombotic agents with at least 500 total patients across all trials or a single trial with more than 200 patients, for being discussed in the full text of the current manuscript); Supplemental table 4 shows investigational agents which did not meet inclusion criteria. | • Studies of pediatric patients |
| • The following characteristics being available<br>• Planned sample size<br>• Defined study intervention<br>• Inclusion and exclusion criteria<br>• Primary endpoint | • Studies which focus on the treatment of diagnosed thrombotic events (rather than prophylactic treatment) |
| COVID-19: Coronavirus Disease 2019, WHO ICTRP: World Health Organization International Clinical Trials Registry Platform |  |

**Supplemental Table 2.**
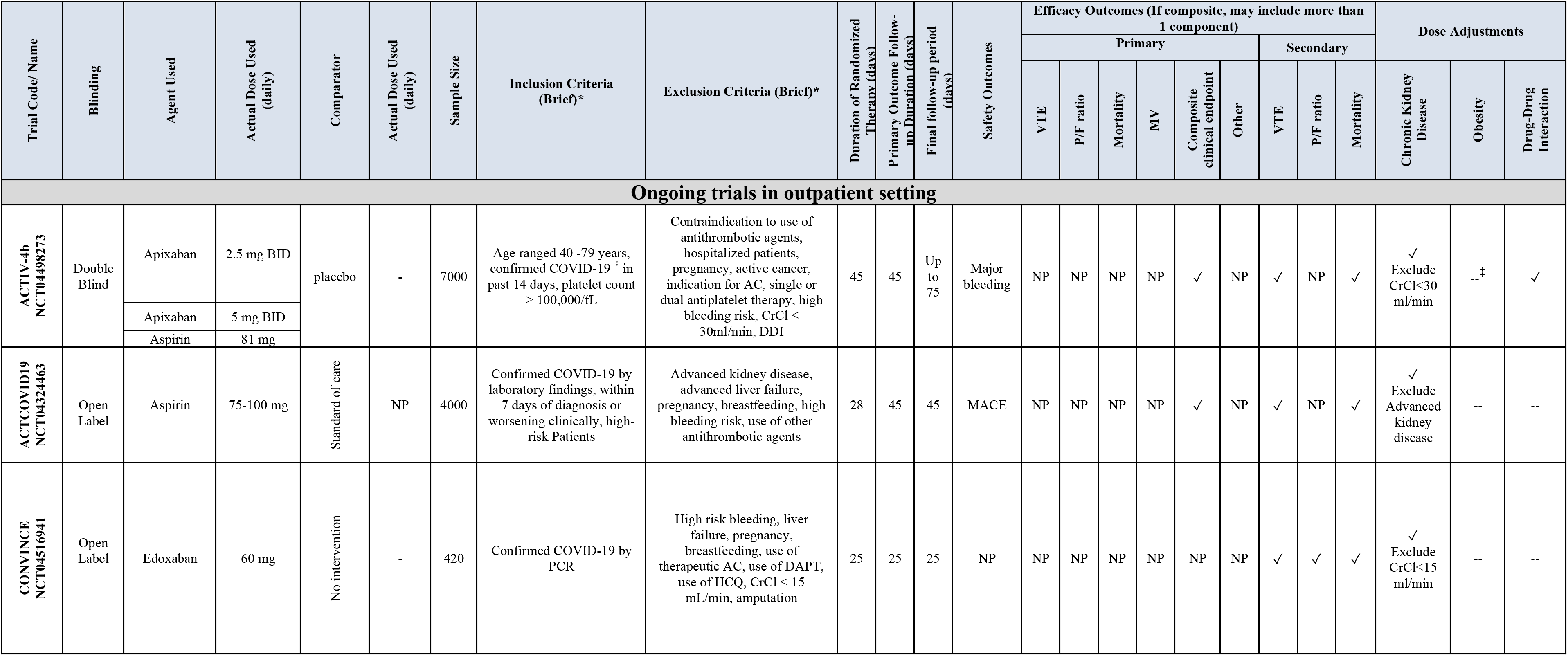
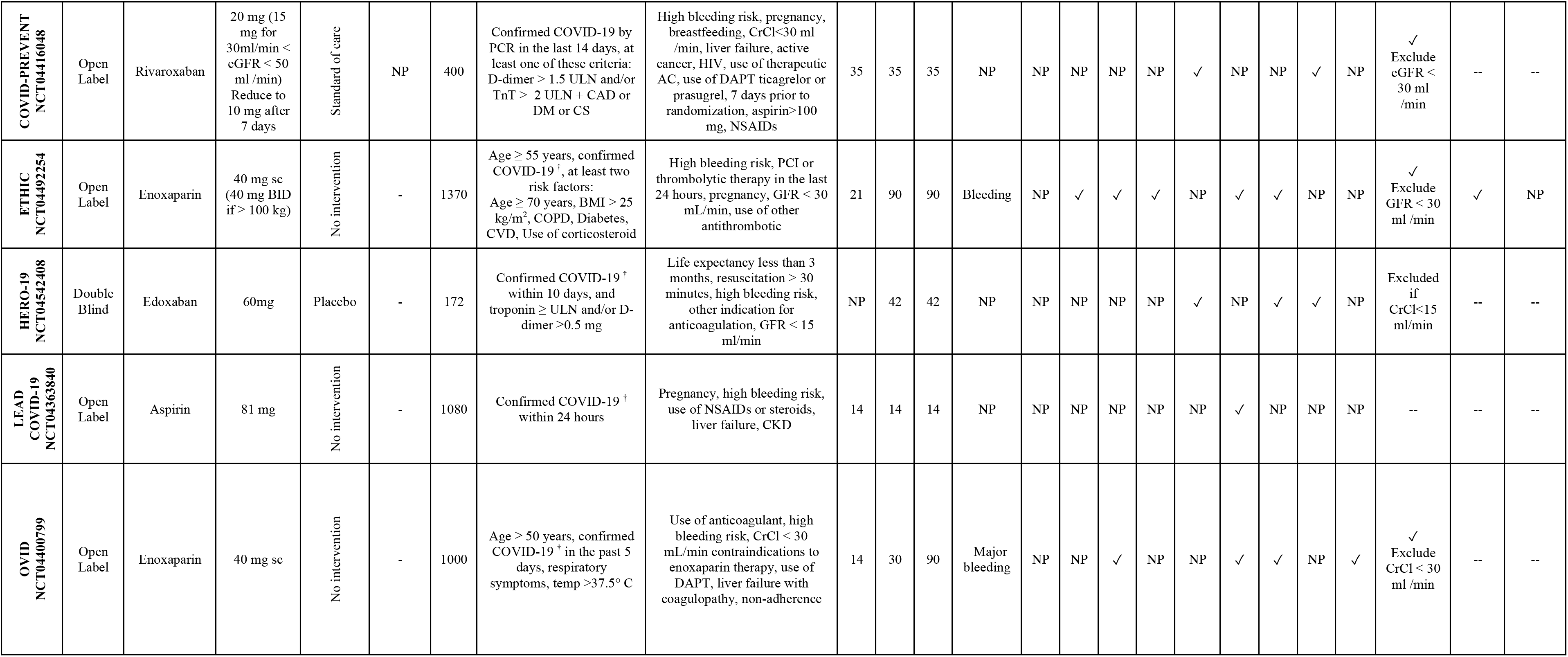
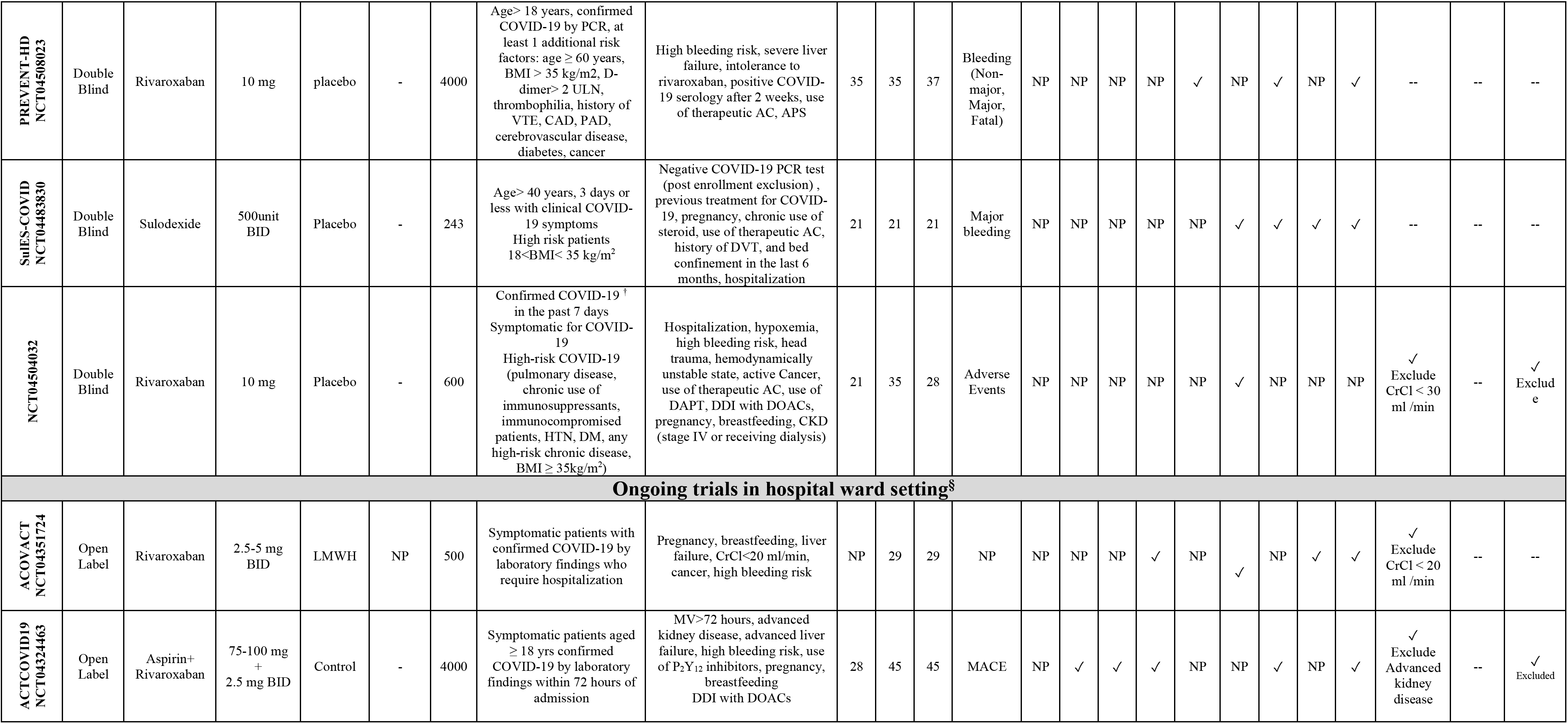
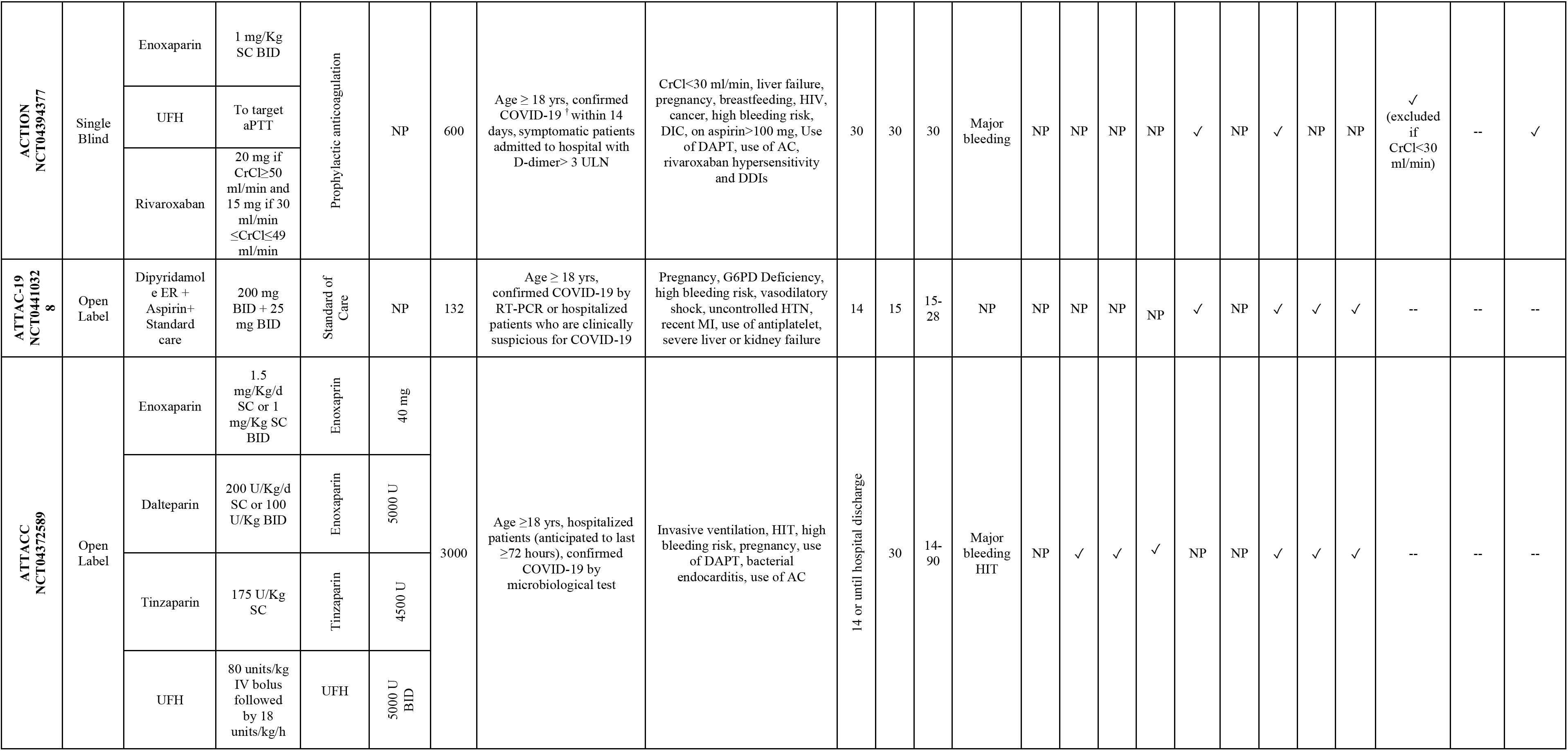
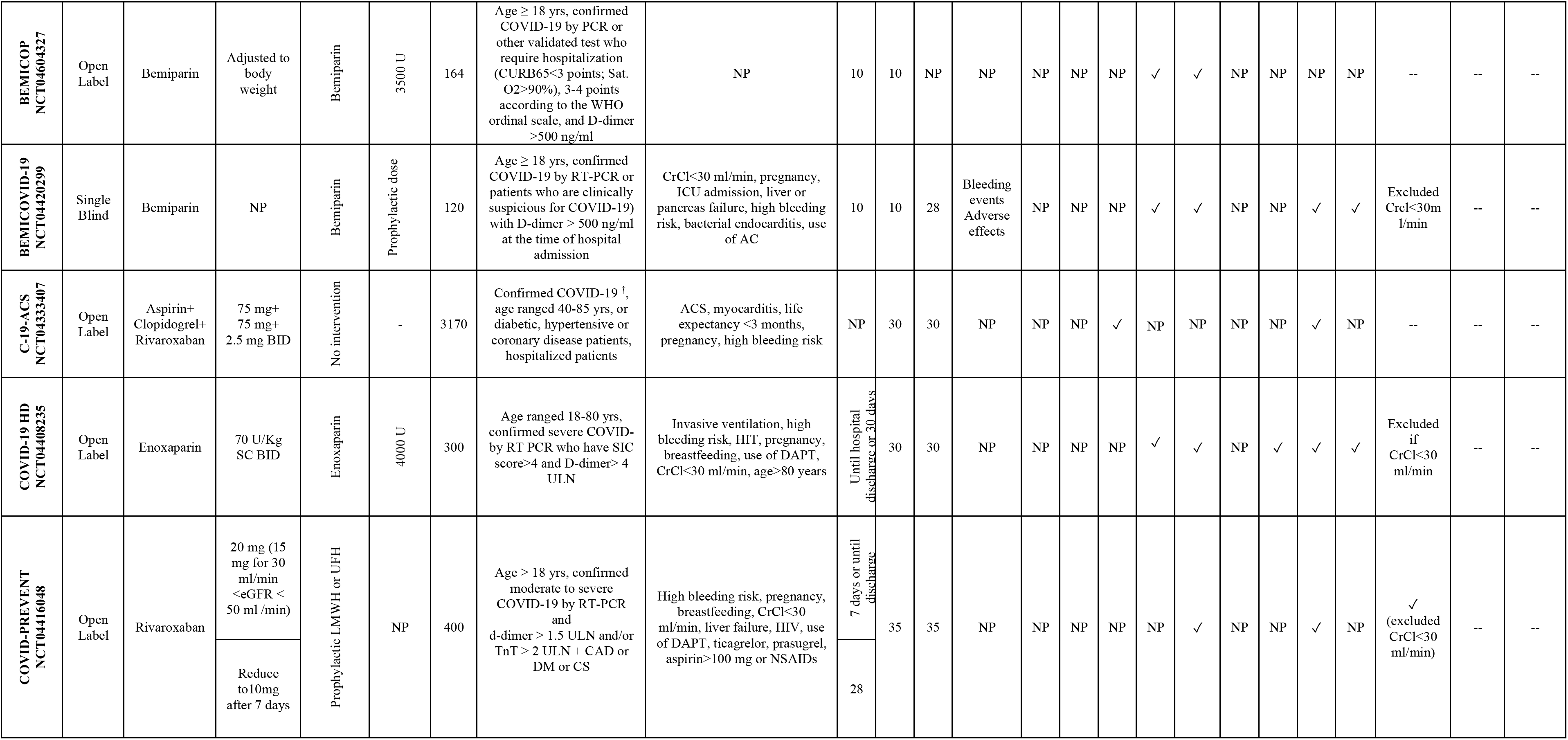
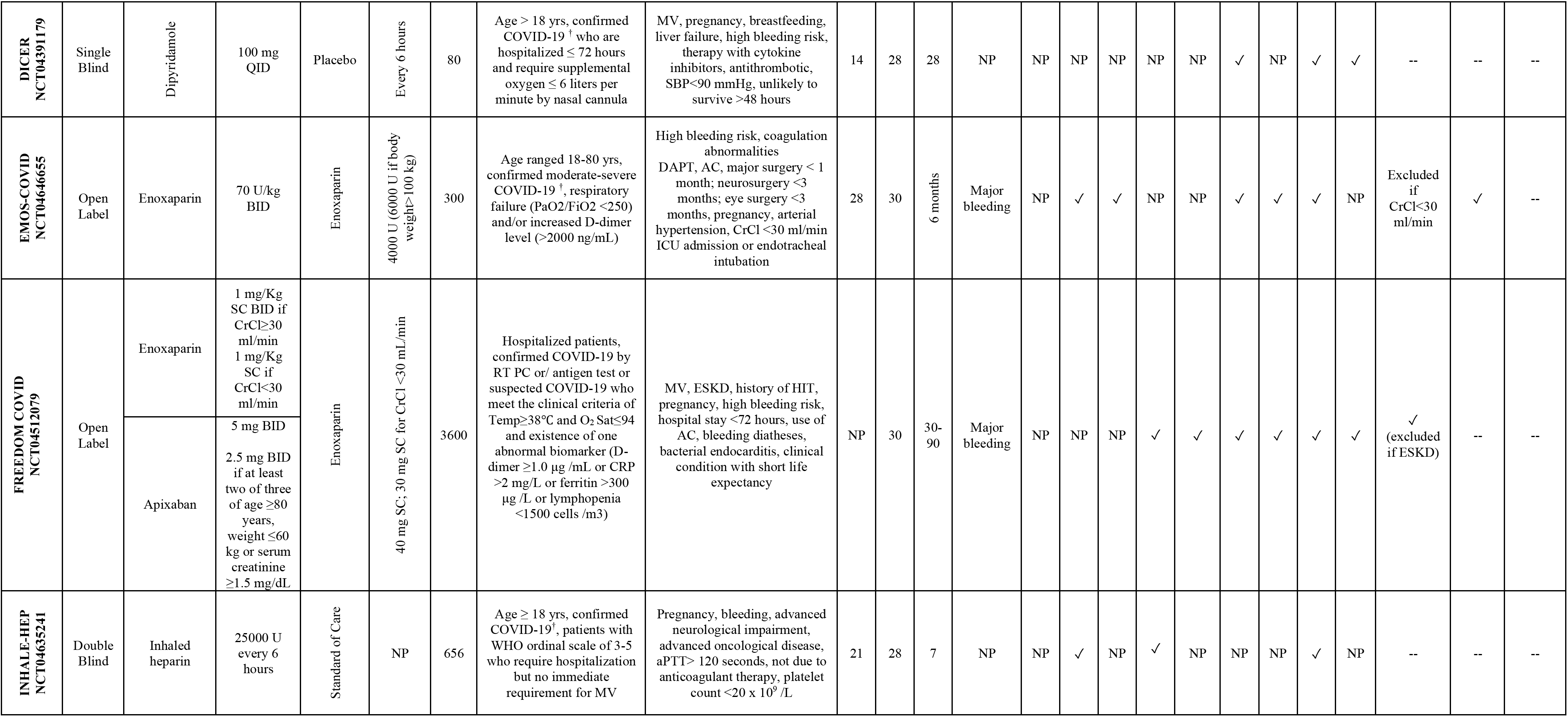
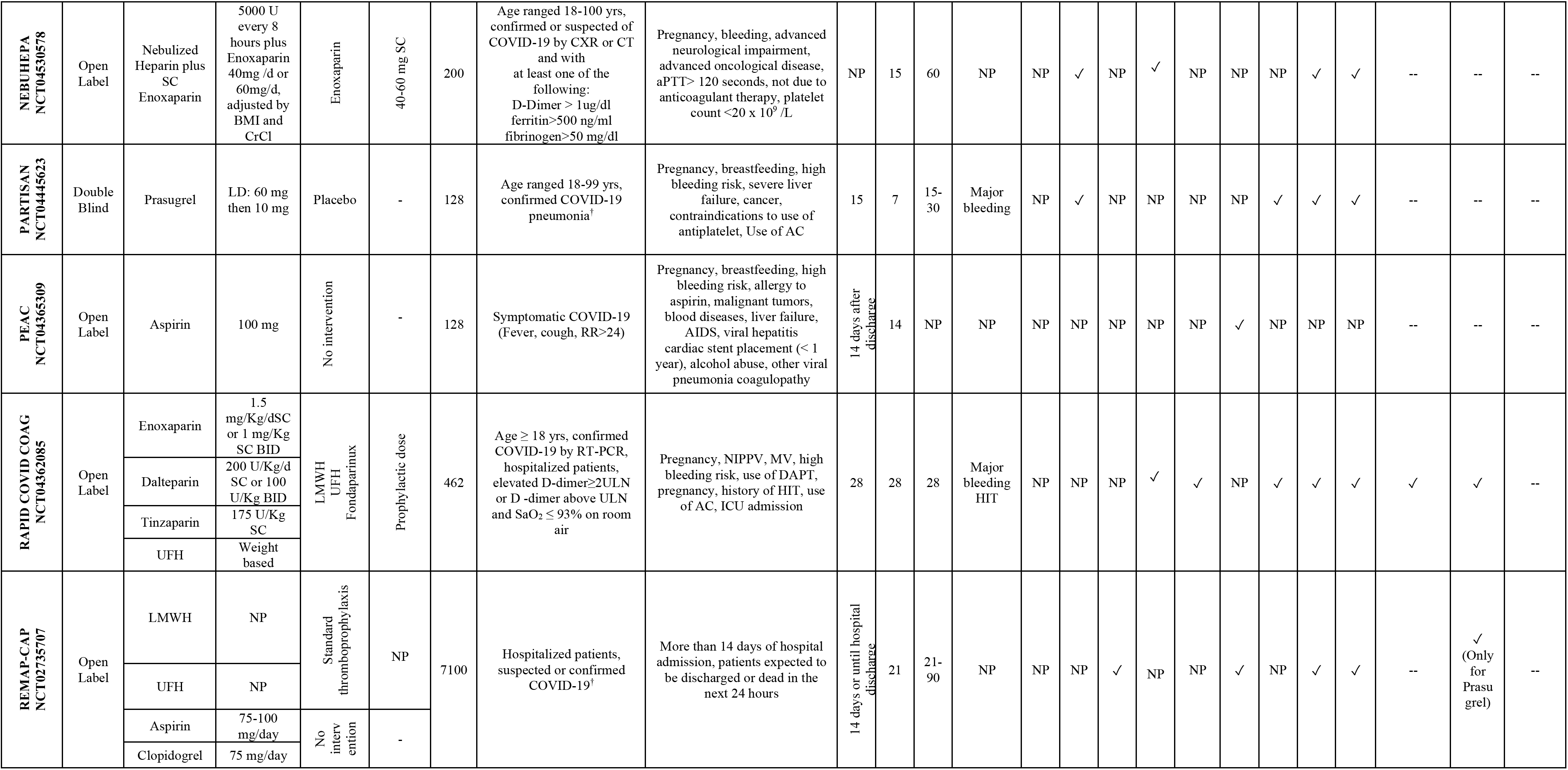
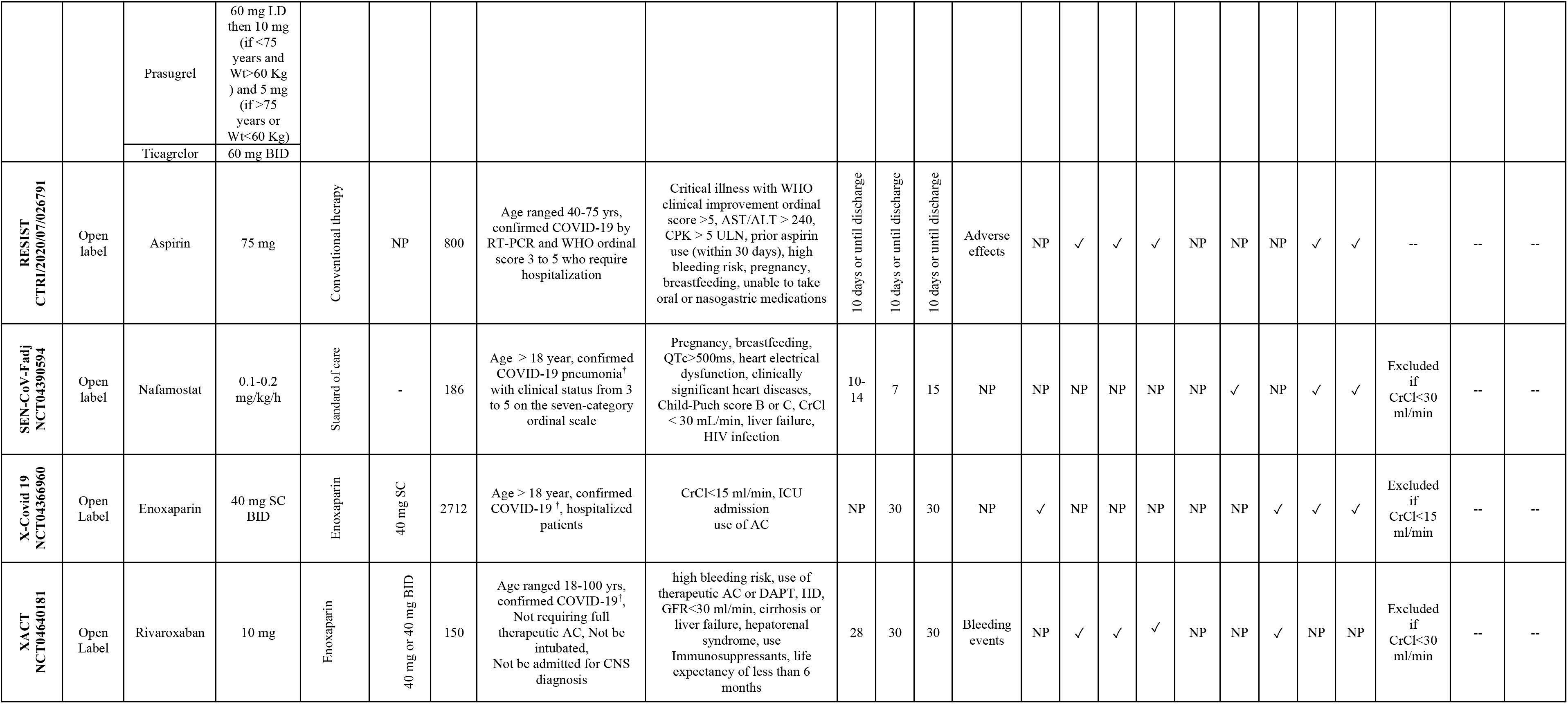
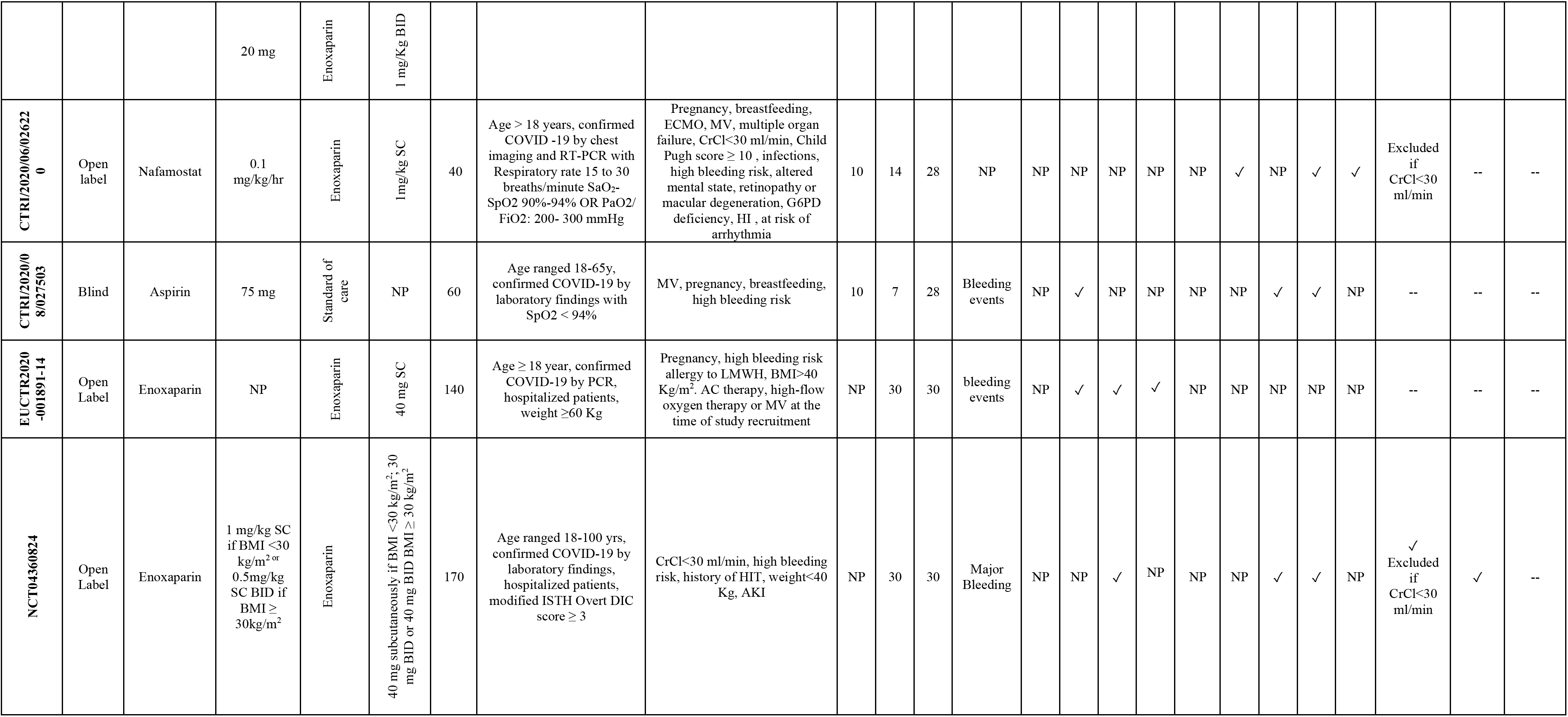
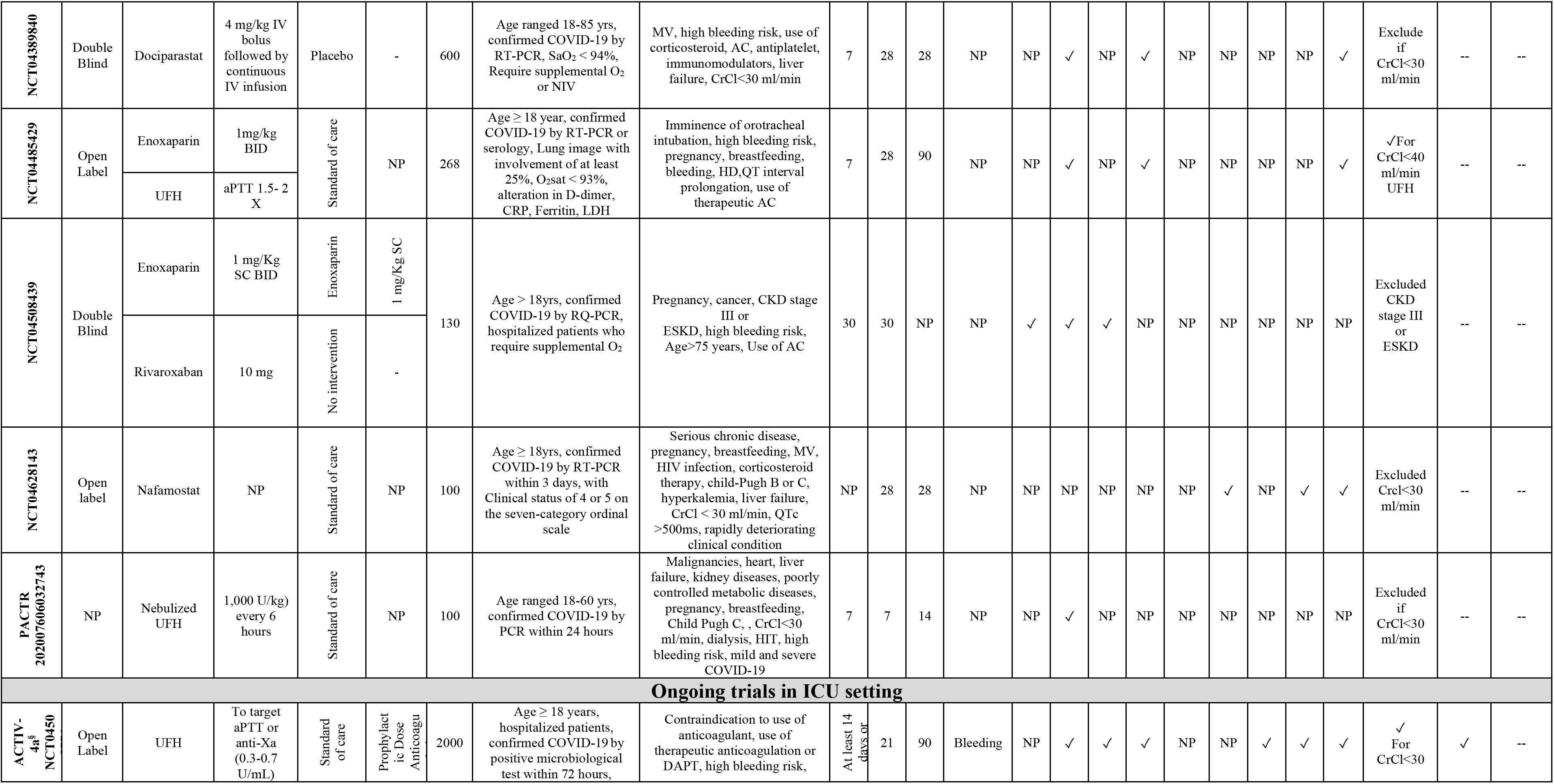
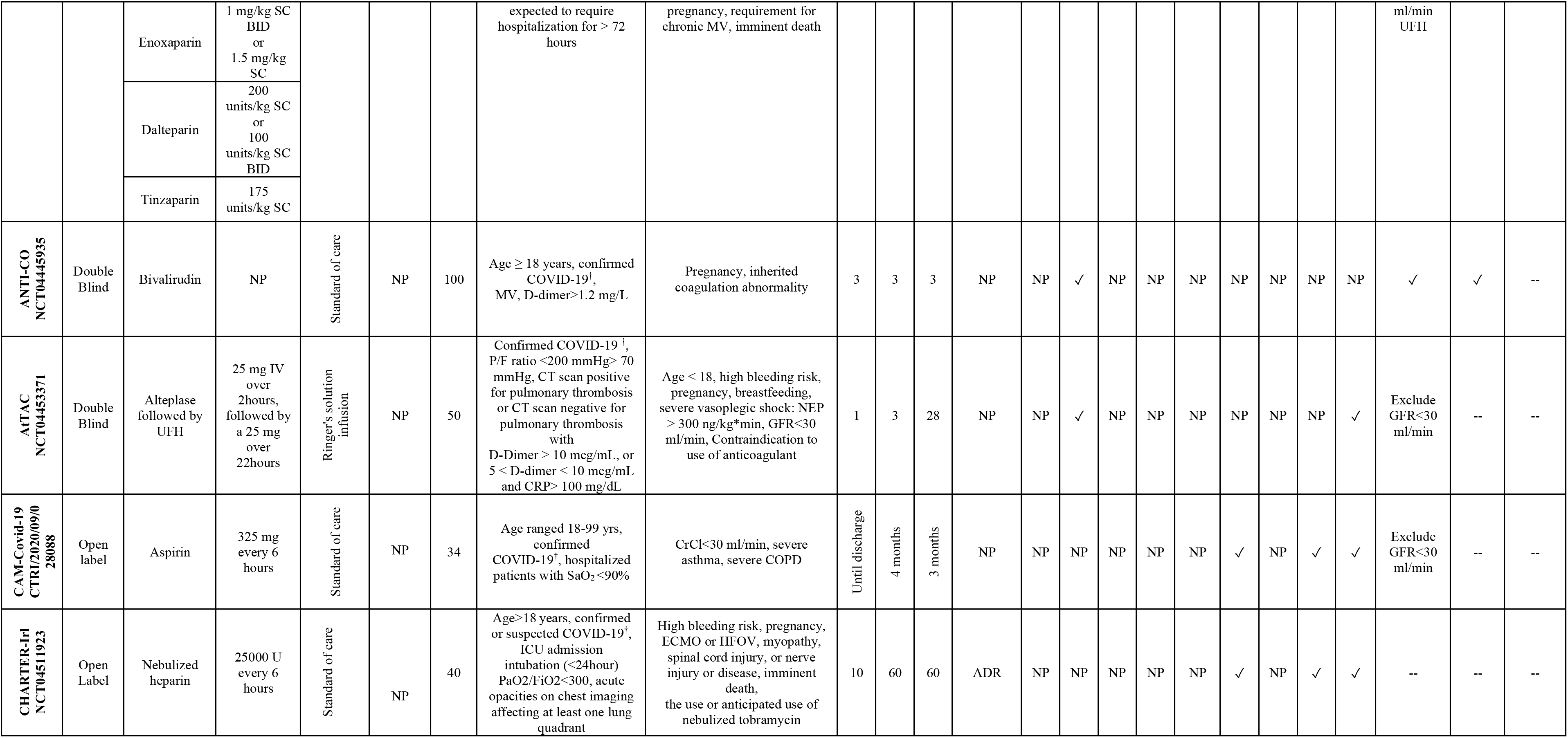
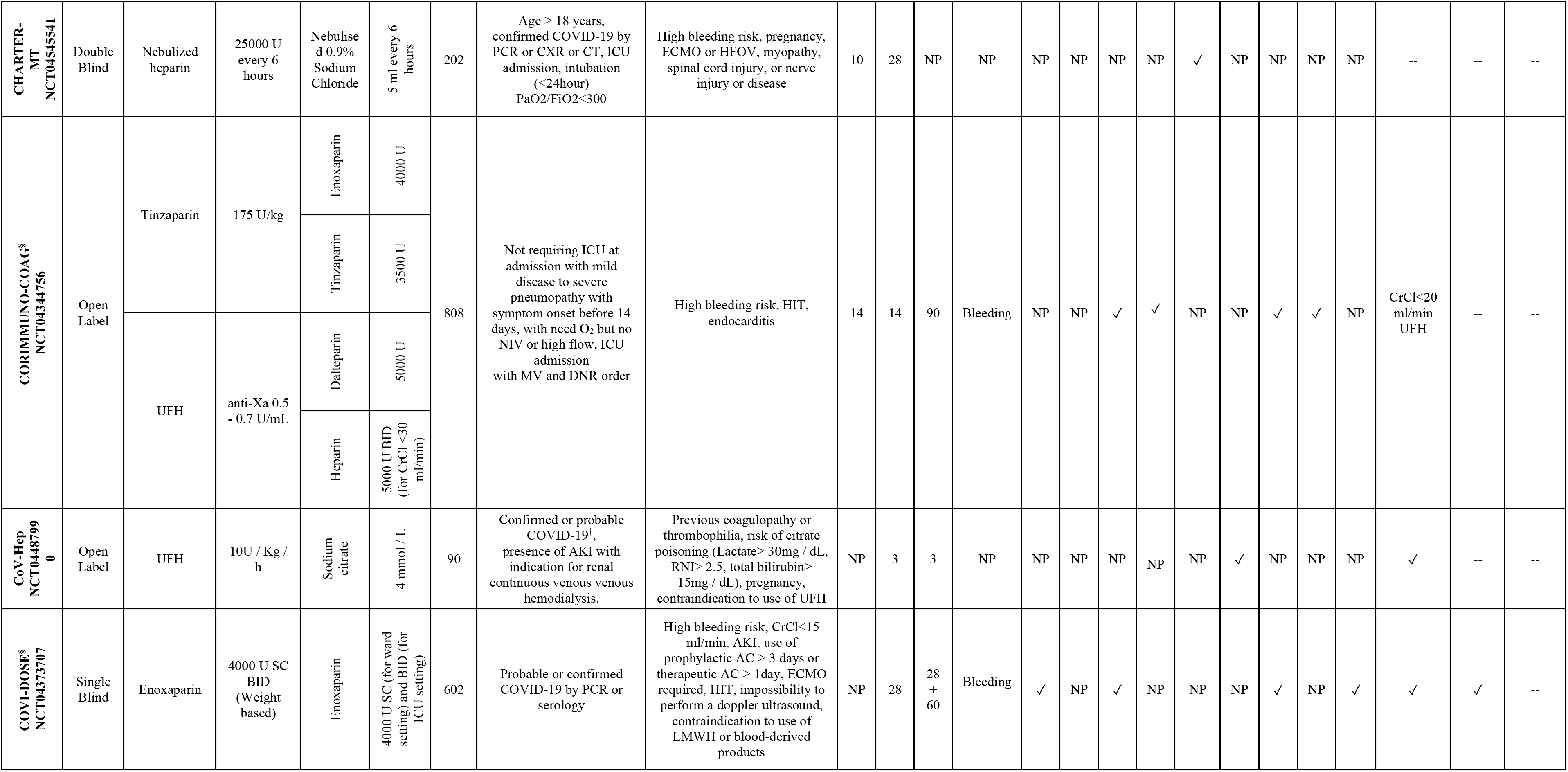
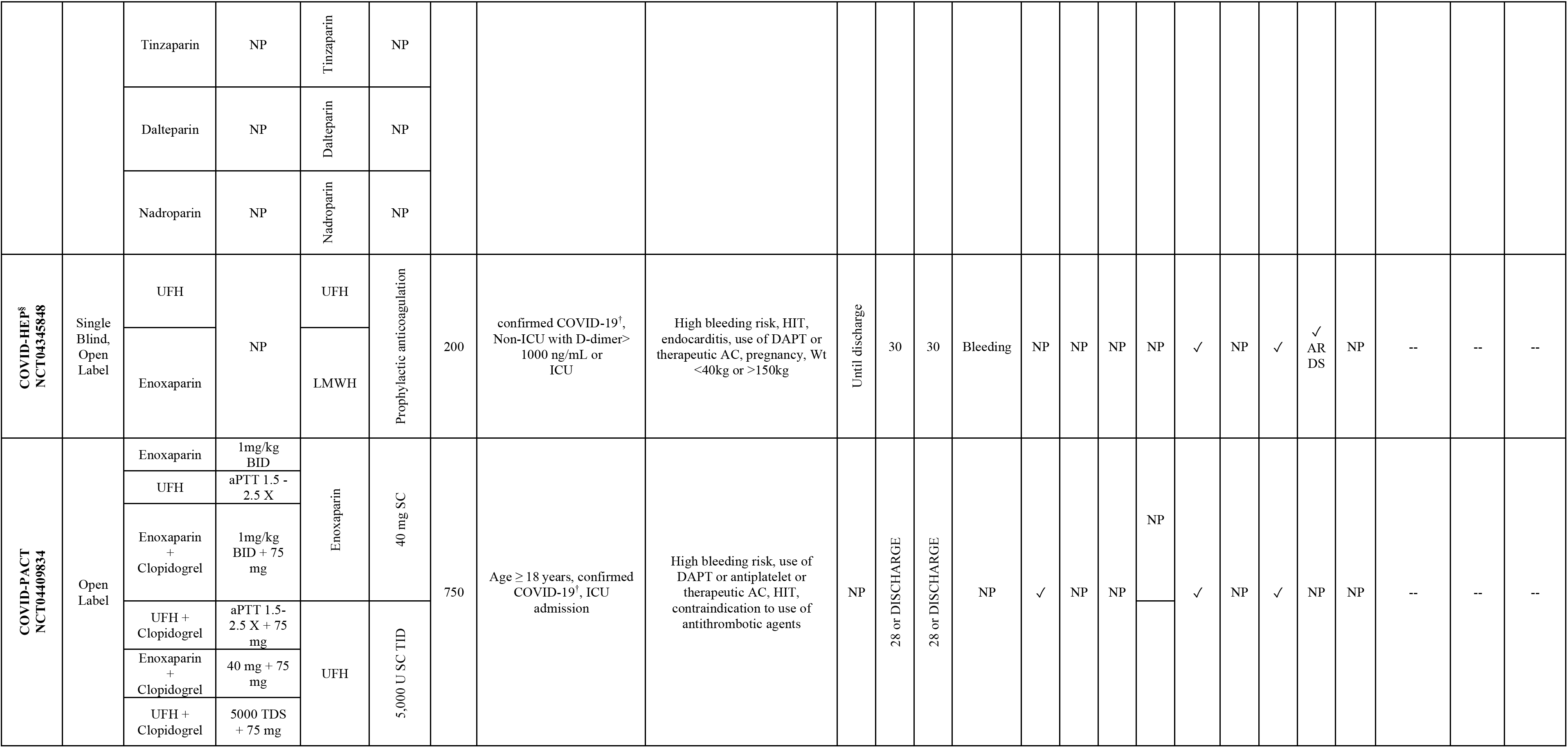
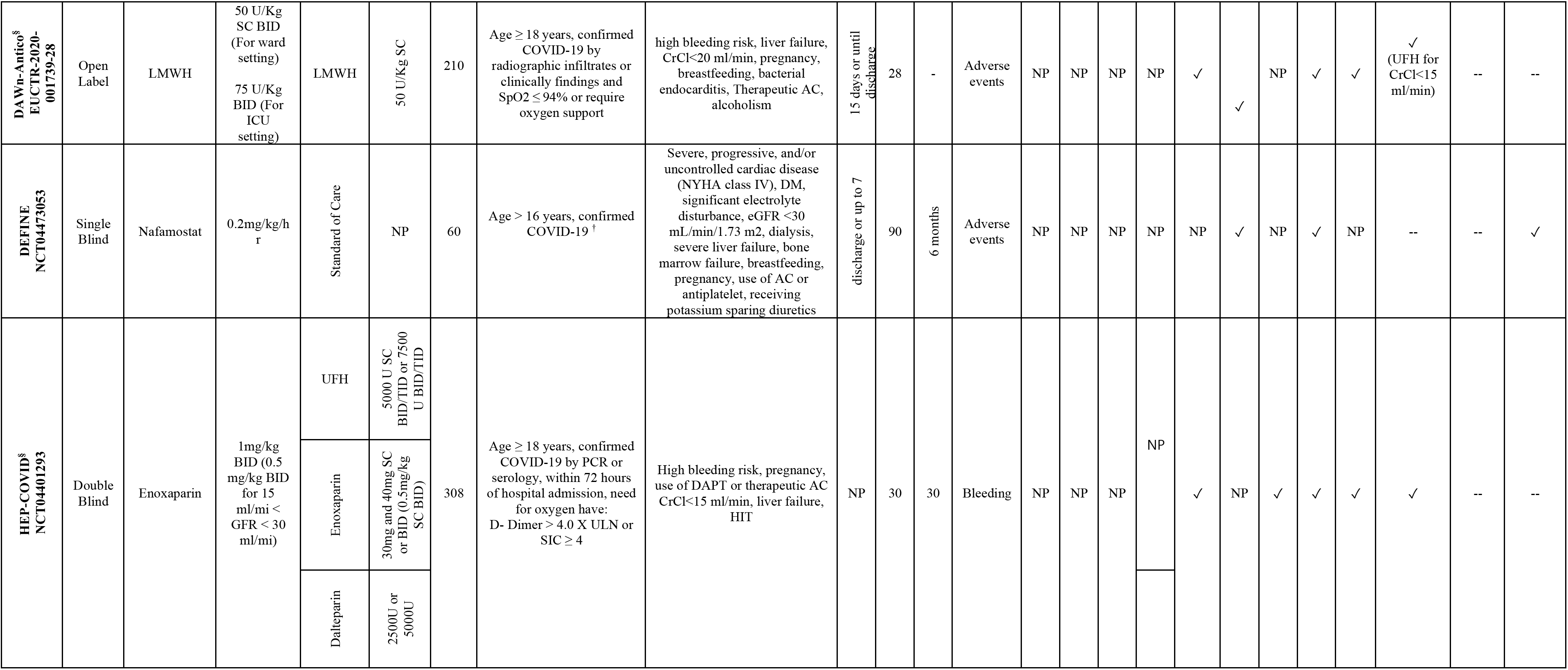
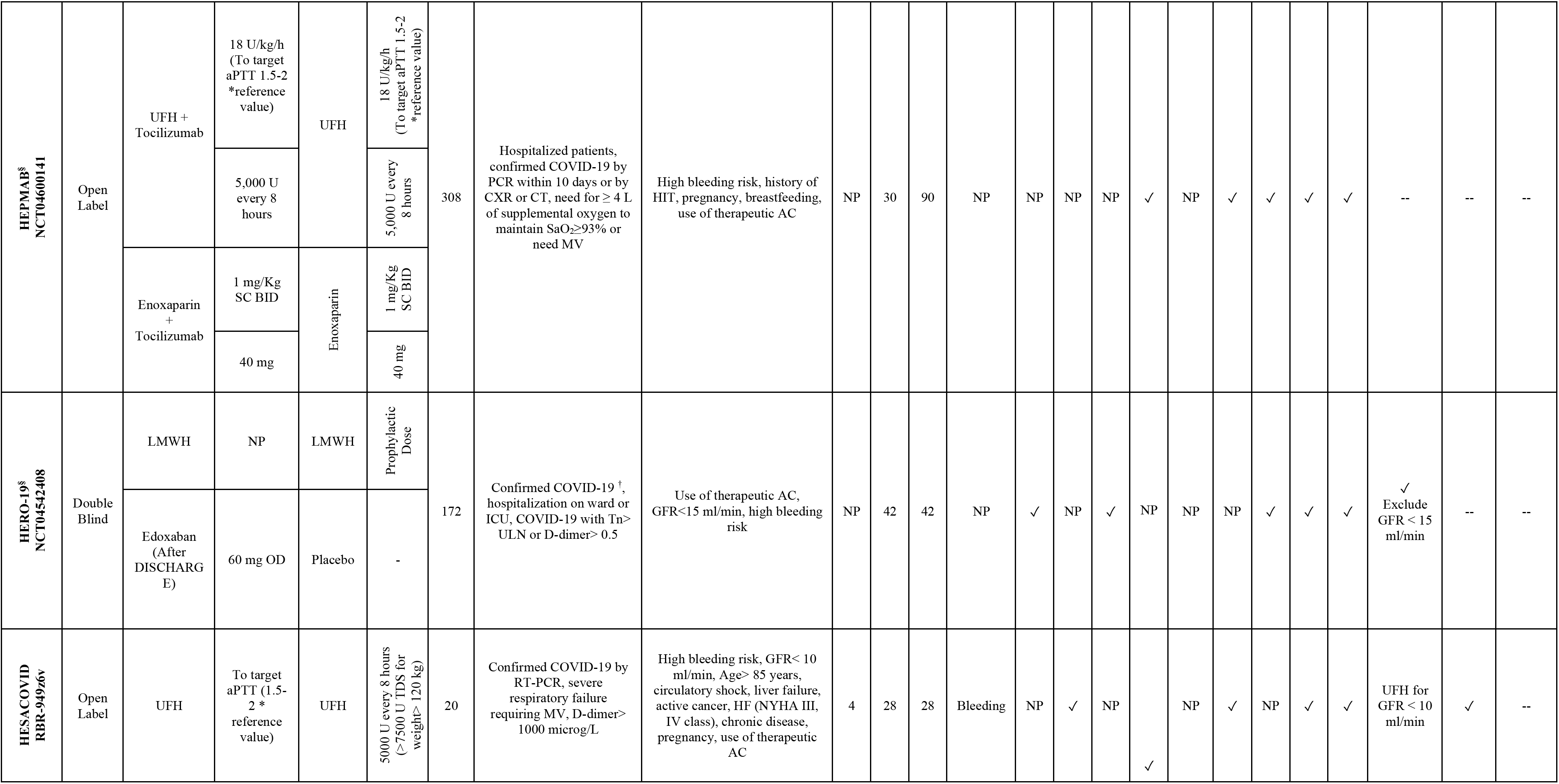
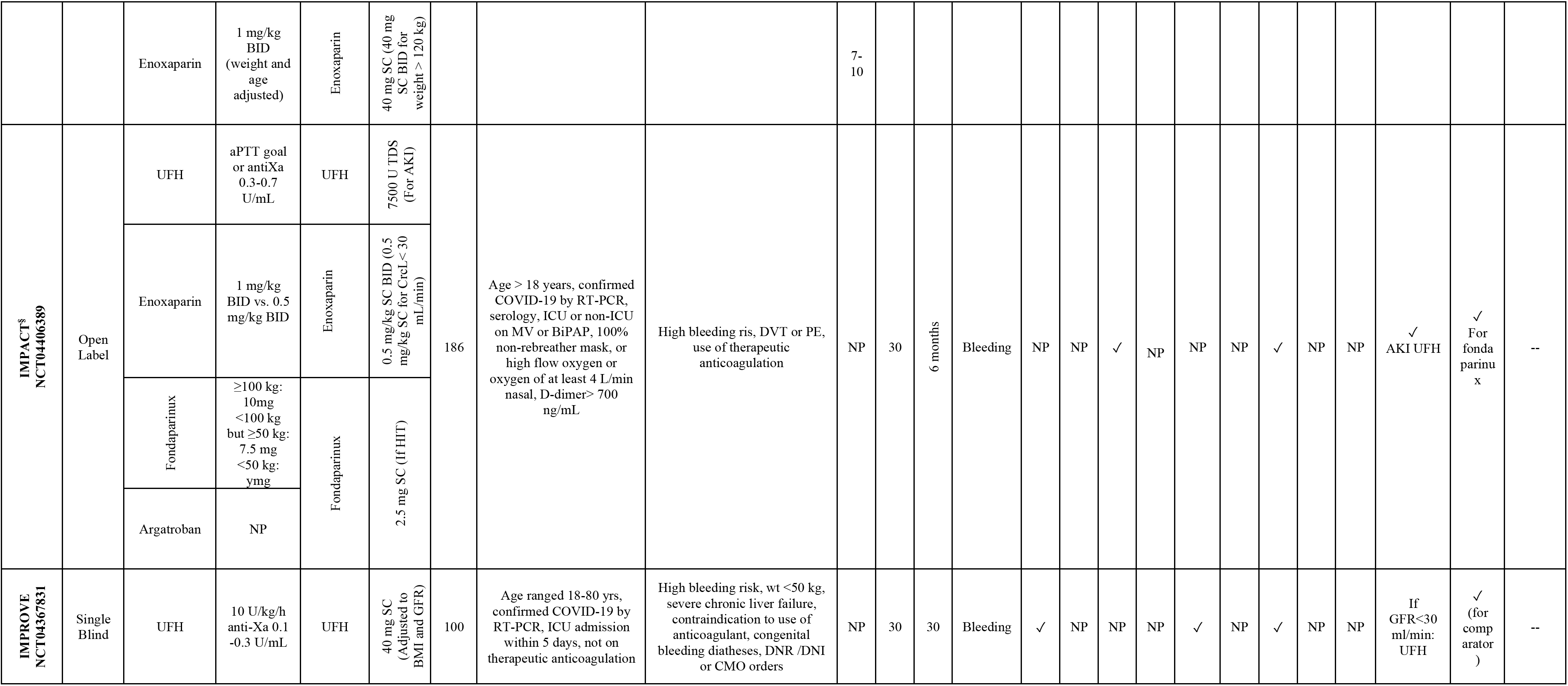
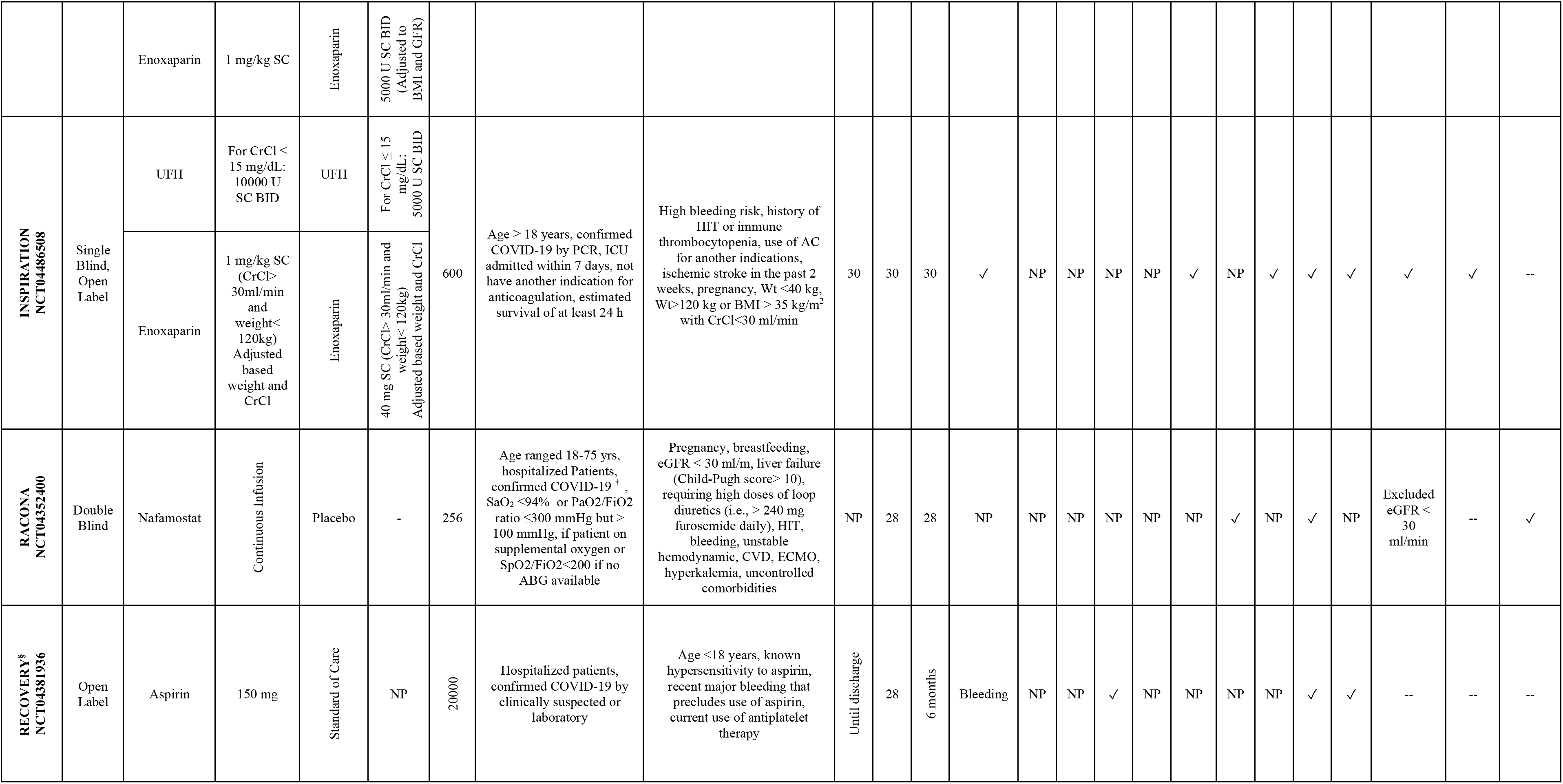
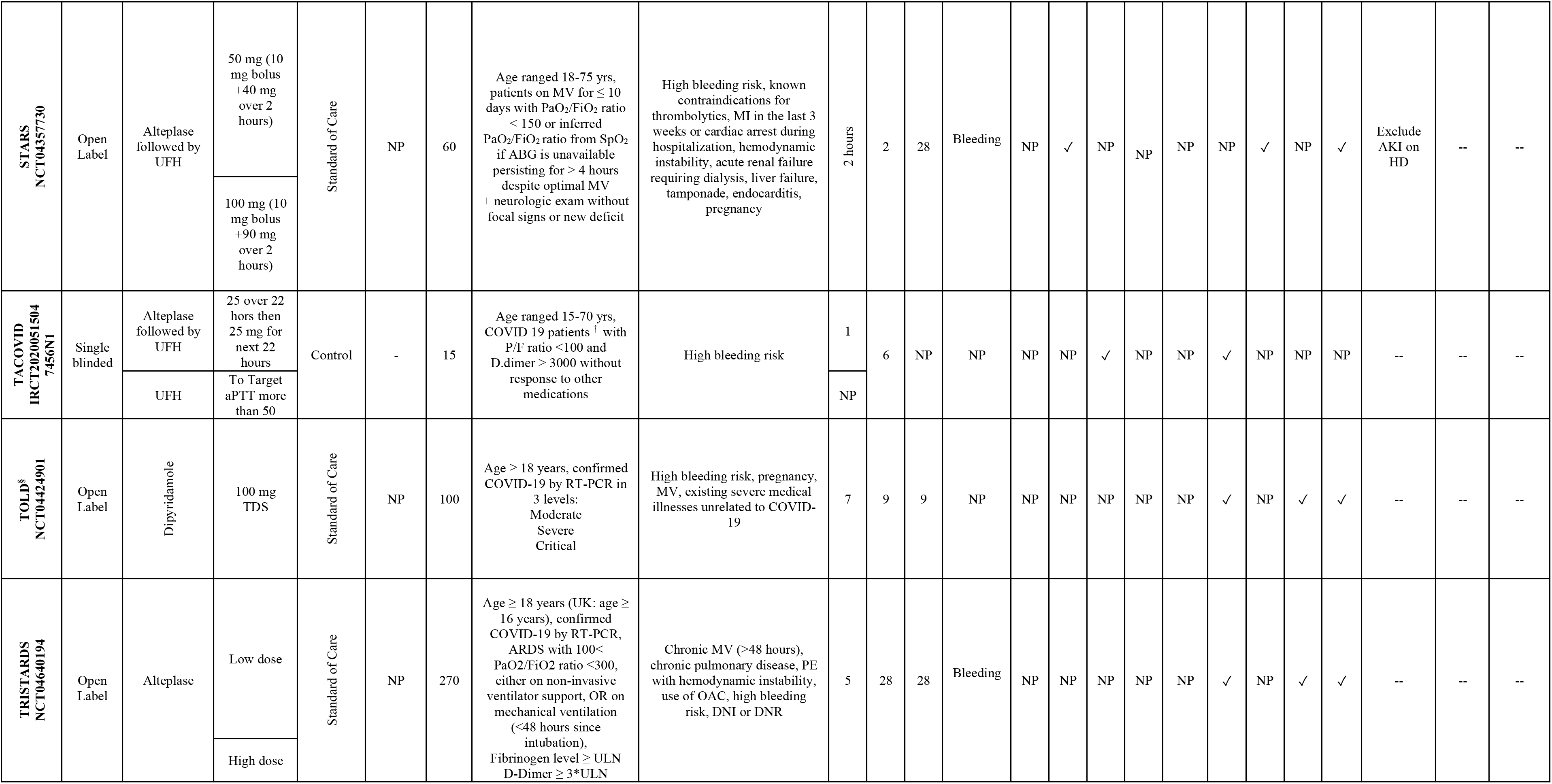
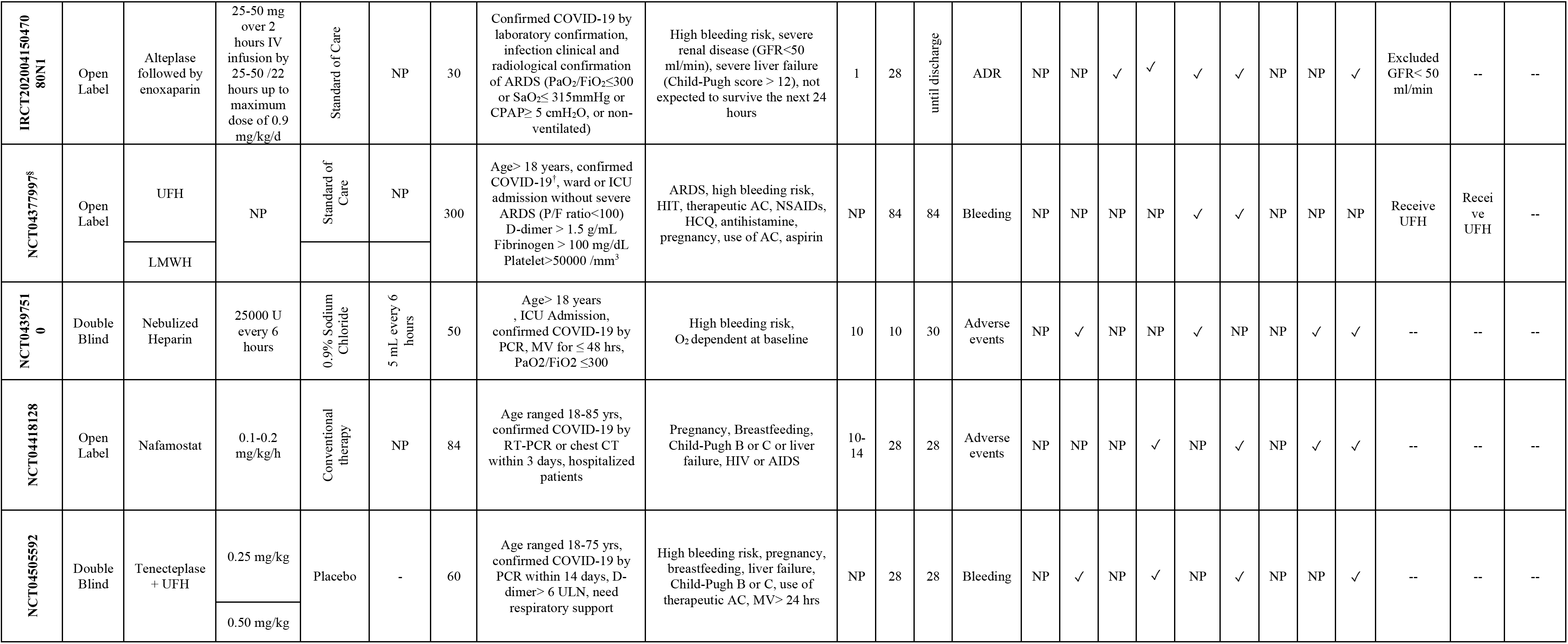
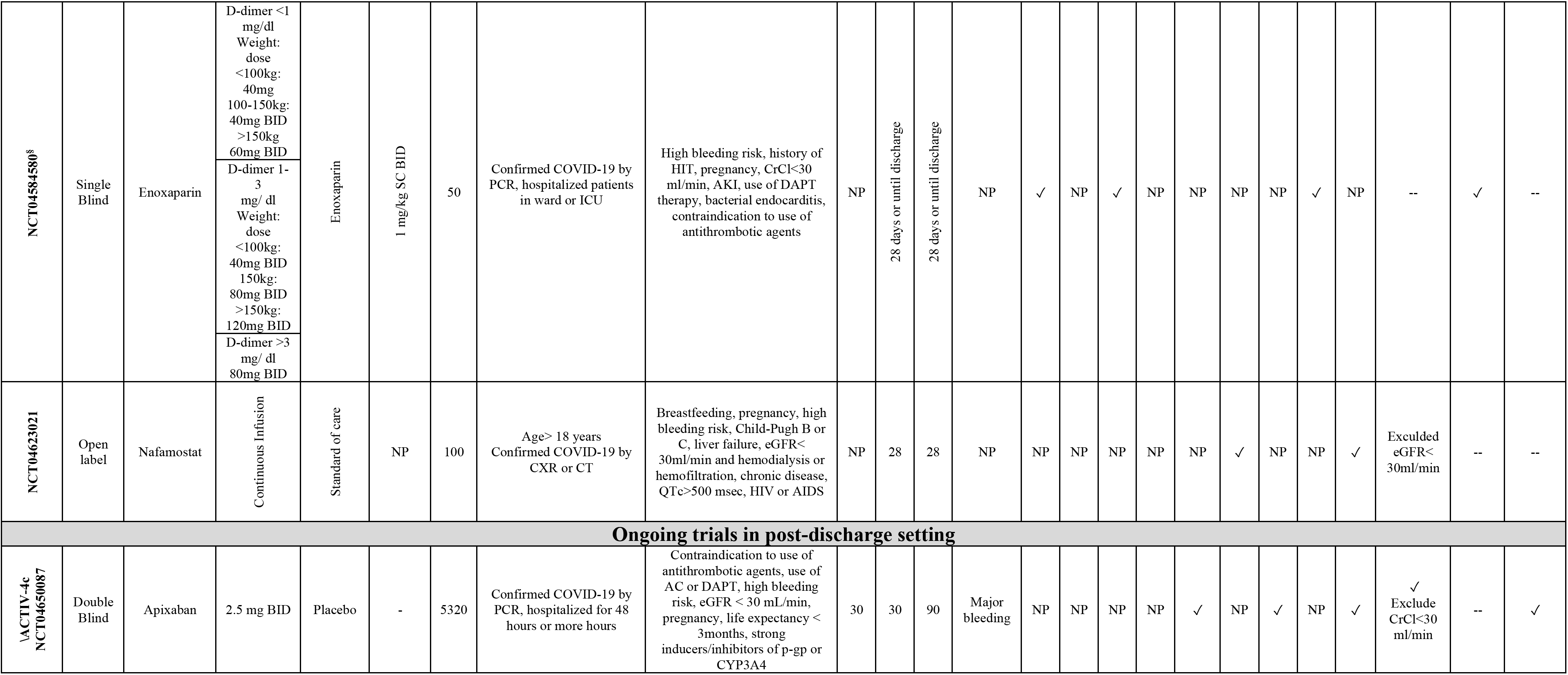
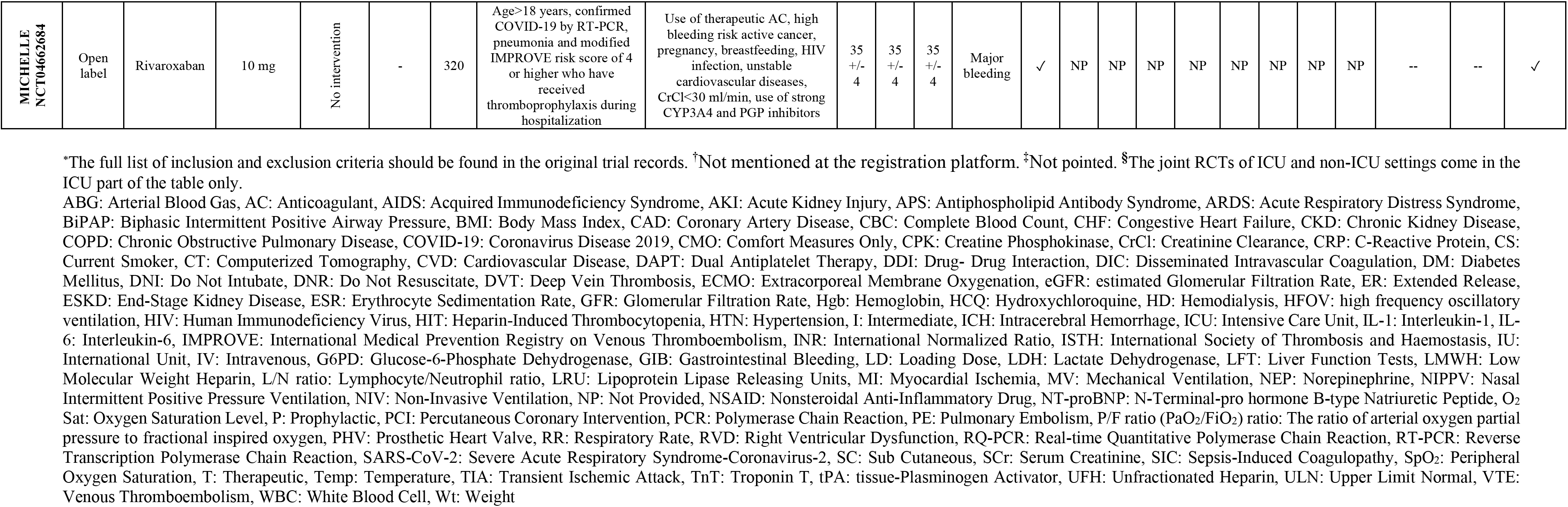
Review of RCTs Categorized Based on Patient Settings.

**Supplemental Table 3.**
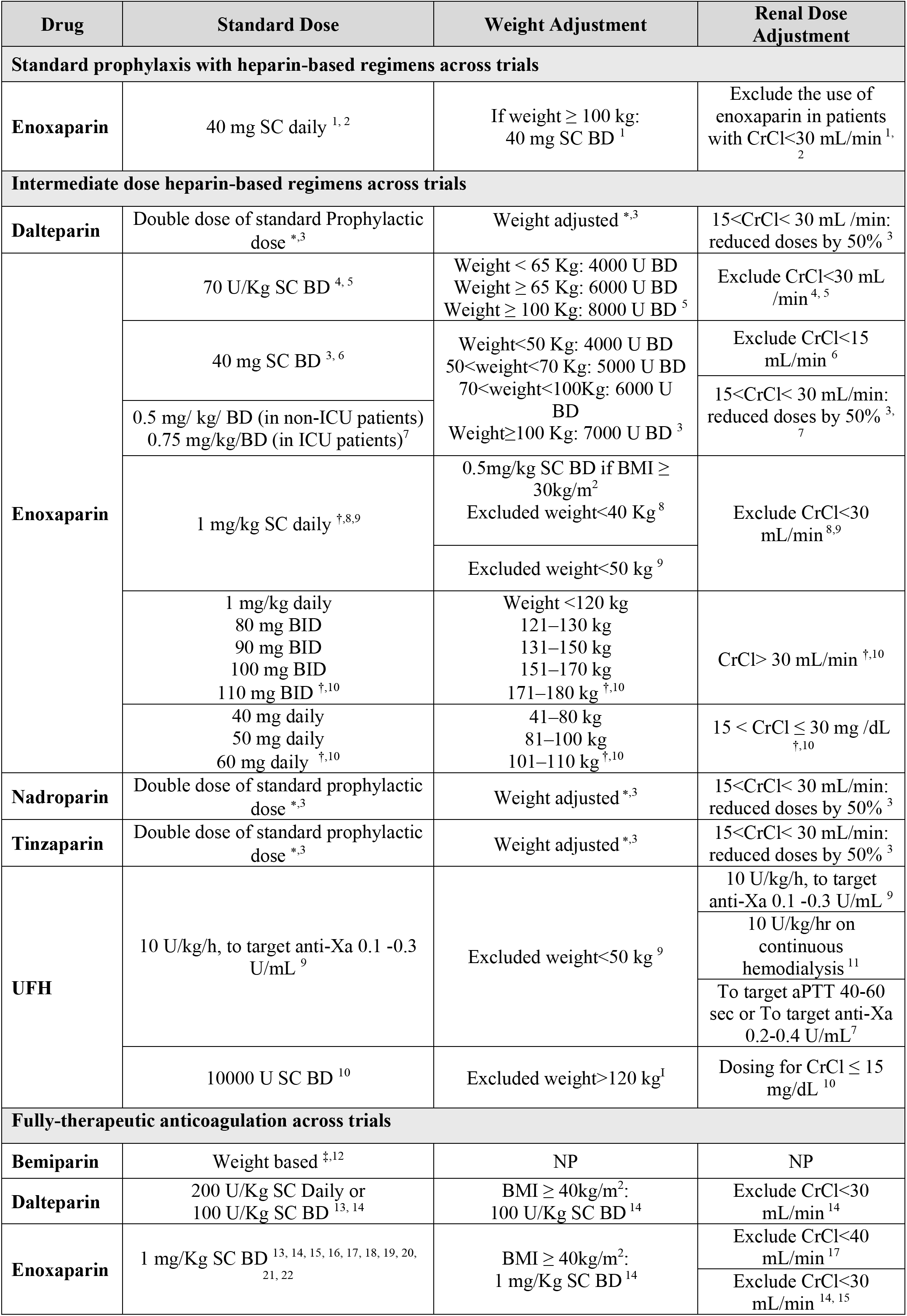
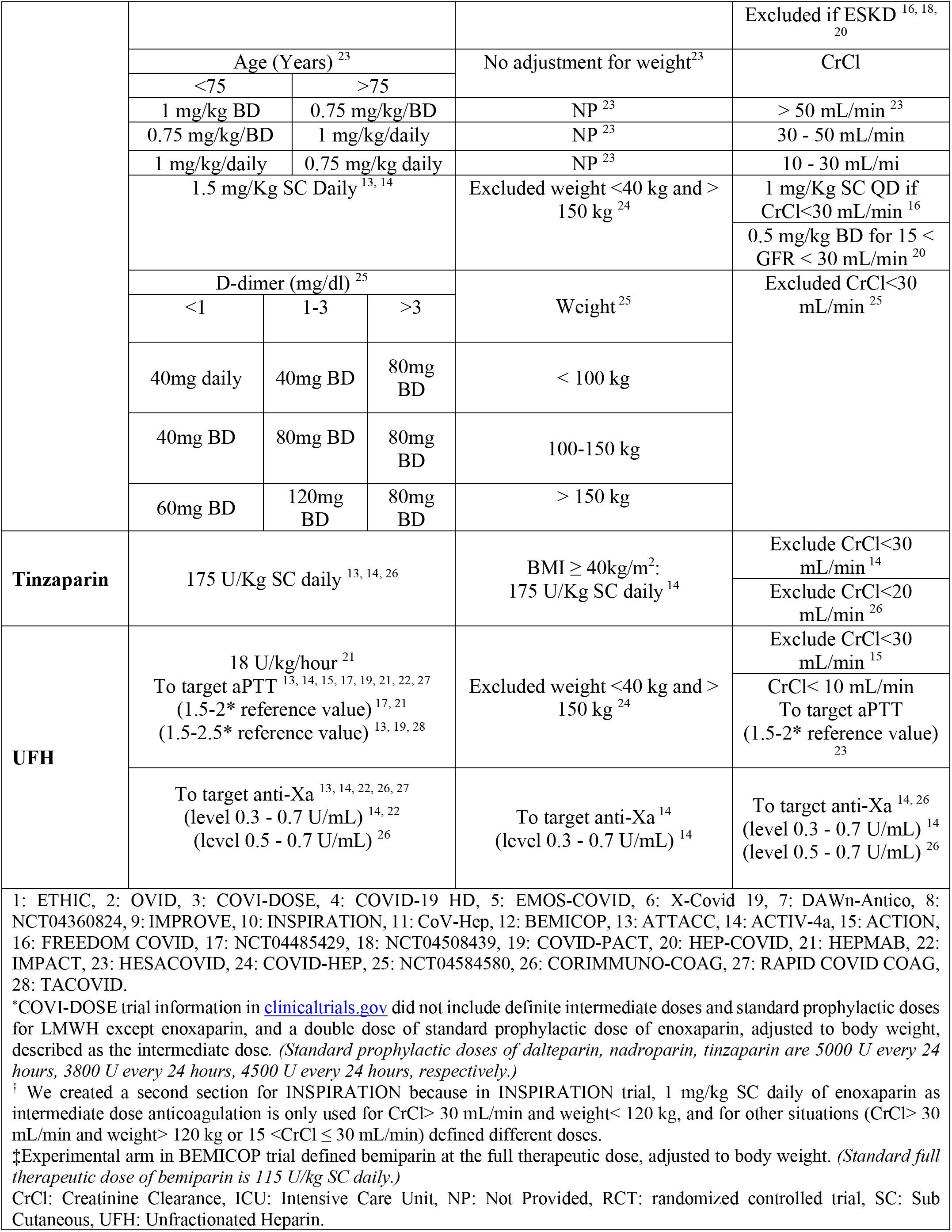
The intensities of heparin derivatives In the Studied RCTs.

**Supplemental Table 4.** Additional Investigational Agents with Antithrombotic Properties Studied in COVID-19.

|  |  |  |  |  |  |
| --- | --- | --- | --- | --- | --- |
| Agent | Mechanism of Action | Ongoing Randomized Trials | Setting | Primary Outcomes Include | Sample Size |
| Crizanlizumab | A monoclonal antibody targeting P-selectin. Crizanlizumab can decrease inflammation by binding to P-selectin, blocking leucocyte and platelet adherence to the vessel wall. | CRITICAL (NCT04435184) | Ward | Level of soluble P-selectin in ng/ml | 50 |
| Defibrotide | Augmenting plasmin activity to hydrolyze fibrin clots; reducing endothelial cell activation, increasing endothelial cell-mediated fibrinolysis, increasing tissue plasminogen activator and thrombomodulin expression; decreasing von Willebrand factor and plasminogen activator inhibitor-1. | DEFACOVID (NCT04348383) | Ward and ICU | All-cause mortality | 120 |
| Recombinant antithrombin III | Inactivation of thrombin, plasmin, and other serine proteases of coagulation (factors IXa, Xa, XIIa). The inactivation of proteases is a major step in the normal clotting process and thrombosis would be caused by active serine proteases if they were not inhibited by antithrombin. | ANTITROMBINA (EUCT2020-001659-42) | Ward | Composite of mortality and need for non-invasive ventilation or mechanical ventilation | 46 |
| Recombinant nematode anticoagulant protein c2 (rNAPc2) | Inhibition of tissue factor/factor VIIa, attenuates coagulation and the interleukin-10 response in human endotoxemia. | ASPEN-COVID-19 | Ward | Change in D-dimer level from baseline, time to recovery within 30 days | 100 |
| Ulinastatin | As a urinary serine protease inhibitor, it inactivates plasmin, elastase, thrombin, factors IXa, Xa, XIa, and XIIa. It also inhibits several pro-inflammatory proteases and decreases inflammatory cytokine. | NCT04393311 | Ward | Time to recovery defined as attaining a score of 6, 7, or 8 on the COVID-19 disease severity scale | 160 |
|  |  | CTRI/2020/06/025704 | Ward | Number of days of use of mechanical ventilation, change from baseline in PaO2/FiO2 ratio | 120 |
|  |  | ChiCTR2000030779 | ICU | Blood gas, SOFA score | 100 |
|  |  | ChiCTR2000032135 | Ward and ICU | Mild cases: the critical illness rate of subjects at weeks 2<br>Severe or critical cases: changes of PaO2/FiO2 from baseline | 50 |
| COVID-19: coronavirus disease 2019; ICU: intensive care unit; SOFA: sequential organ failure assessment. |  |  |  |  |  |

## References

1. Piazza G, Campia U, Hurwitz S, et al. Registry of arterial and venous thromboembolic complications in patients with COVID-19. J Am Coll Cardiol. 2020;76:2060–72.

2. Nadkarni GN, Lala A, Bagiella E, et al. Anticoagulation, mortality, bleeding and pathology among patients hospitalized with COVID-19: a single health system study. J Am Coll Cardiol. 2020;76:1815–26.

3. Schulman S, Hu Y, Konstantinides S. Venous Thromboembolism in COVID-19. J. Thromb. Haemost. 2020[E-pub ahead of print];DOI https://doi.org/10.1055/s-0040-1718532.

4. Voicu S, Bonnin P, Stépanian A, et al. High prevalence of deep vein thrombosis in mechanically ventilated COVID-19 patients. J. Am. Coll. Cardiol. 2020[E-pub ahead of print];https://doi.org/10.1016/j.jacc.2020.05.053.

5. Nopp S, Moik F, Jilma B, Pabinger I, Ay C. Risk of venous thromboembolism in patients with COVID-19: A systematic review and meta-analysis. Res Prac Thromb Haemost. 2020;6:1178–91.

6. Jiménez D, García-Sanchez A, Rali P, et al. Incidence of venous thromboembolism and bleeding among hospitalized patients with COVID-19: a systematic review and meta-analysis. CHEST. 2020 [E-pub ahead of print];https://doi.org/10.1016/j.chest.2020.11.005.

7. Ackermann M, Verleden SE, Kuehnel M, et al. Pulmonary vascular endothelialitis, thrombosis, and angiogenesis in Covid-19. N Engl J Med. 2020;383:120–8.

8. Wichmann D, Sperhake J-P, Lütgehetmann M, et al. Autopsy findings and venous thromboembolism in patients with COVID-19: a prospective cohort study. Ann Intern Med. 2020;173:268–77.

9. Fernández-Capitán C, Barba R, del Carmen Díaz-Pedroche M, et al. Presenting Characteristics, Treatment Patterns, and Outcomes among Patients with Venous Thromboembolism during Hospitalization for COVID-19. Semin. Thromb. Hemost. 2020[E-pub ahead of print];https://doi.org/10.1055/s-0040-1718402.

10. Libby P, Lüscher T. COVID-19 is, in the end, an endothelial disease. Eur Heart J. 2020;41:3038–44.

11. Giustino G, Pinney SP, Lala A, et al. Coronavirus and Cardiovascular Disease, Myocardial Injury, and Arrhythmia: JACC Focus Seminar. J Am Coll Cardiol. 2020;76:2011–23.

12. Skendros P, Mitsios A, Chrysanthopoulou A, et al. Complement and tissue factor–enriched neutrophil extracellular traps are key drivers in COVID-19 immunothrombosis. J Clin Investig. 2020;130:6151–7.

13. Connors JM, Levy JH. COVID-19 and its implications for thrombosis and anticoagulation. Blood. 2020;135:2033–40.

14. Koupenova M. Potential Role of Platelets in COVID-19: Implications for Thrombosis. Res Prac Thromb Haemost. 2020;4:737–40.

15. Siddiqi HK, Libby P, Ridker PM. COVID-19–A vascular disease. Trends Cardiovasc. Med. 2020;https://doi.org/10.1016/j.tcm.2020.10.005.

16. Stefely JA, Christensen BB, Gogakos T, et al. Marked factor V activity elevation in severe COVID-19 is associated with venous thromboembolism. Am J Hematol. 2020;95:1522–30.

17. Becker RC. COVID-19 update: Covid-19-associated coagulopathy. J Thromb Thrombolys. 2020;50:54–67.

18. Zuo Y, Zuo M, Yalavarthi S, et al. Neutrophil extracellular traps and thrombosis in COVID-19. J Thromb Thrombolys. 2020;5:e138999.

19. Singhania N, Bansal S, Nimmatoori DP, Ejaz AA, McCullough PA, Singhania G. Current overview on hypercoagulability in COVID-19. Am J Cardiovasc Drugs. 2020;20:393–403.

20. Jin S, Jin Y, Xu B, Hong J, Yang X. Prevalence and impact of coagulation dysfunction in COVID-19 in China: a meta-analysis. Thromb Haemost. 2020;120:1524–35.

21. Katneni UK, Alexaki A, Hunt RC, et al. Coagulopathy and thrombosis as a result of severe COVID-19 infection: a microvascular focus. Thromb Haemost. 2020[E-pub ahead of print];DOI:https://doi.org/10.1055/s-0040-1715841.

22. Marchandot B, Trimaille A, Curtiaud A, et al. Staging severity of COVID-19 according to hemostatic abnormalities (CAHA Score). Thromb Haemost. 2020[E-pub ahead of print];DOI https://doi.org/10.1055/s-0040-1715836.

23. Zuo Y, Estes SK, Ali RA, et al. Prothrombotic autoantibodies in serum from patients hospitalized with COVID-19. Sci. Transl. Med. 2020[E-pub ahead of print];DOI: 10.1126/scitranslmed.abd3876.

24. Borghi MO, Beltagy A, Garrafa E, et al. Anti-phospholipid antibodies in COVID-19 are different from those detectable in the anti-phospholipid syndrome. Frontiers in immunology. 2020;11:2692–9.

25. Szekely Y, Lichter Y, Taieb P, et al. The Spectrum of Cardiac Manifestations in Coronavirus Disease 2019 (COVID-19)-a Systematic Echocardiographic Study. Circulation. 2020;142:342–53.

26. Giustino G, Croft LB, Stefanini GG, et al. Characterization of myocardial injury in patients with COVID-19. J Am Coll Cardiol. 2020;76:2043–55.

27. Moores LK, Tritschler T, Brosnahan S, et al. Prevention, Diagnosis, and Treatment of VTE in Patients With Coronavirus Disease 2019: CHEST Guideline and Expert Panel Report. Chest. 2020;158:1143–63.

28. Andreini D, Arbelo E, Barbato E, et al. ESC guidance for the diagnosis and management of CV disease during the COVID-19 pandemic. ESC. June 2020. Available at: https://www.escardio.org/Education/COVID-19-and-Cardiology/ESC-COVID-19-Guidance. Accessed October 1, 2020.

29. The COVID-19 Treatment Guidelines Panel’s Statement on the Emergency Use Authorization of Convalescent Plasma for the Treatment of COVID-19. National Institutes of Health. September 2020. Available at: https://www.covid19treatmentguidelines.nih.gov/. Accessed Ocotober 1,2020.

30. Thachil J, Tang N, Gando S, et al. ISTH interim guidance on recognition and management of coagulopathy in COVID-19. J Thromb Haemost. 2020;18:1023–6.

31. Bikdeli B, Madhavan MV, Jimenez D, et al. COVID-19 and Thrombotic or Thromboembolic Disease: Implications for Prevention, Antithrombotic Therapy, and Follow-Up: JACC State-of-the-Art Review. J Am Coll Cardiol. 2020;75:2950–73.

32. Llitjos JF, Leclerc M, Chochois C, et al. High incidence of venous thromboembolic events in anticoagulated severe COVID-19 patients. J Thromb Haemost. 2020;18:1743–6.

33. Al-Samkari H, Karp Leaf RS, Dzik WH, et al. COVID and Coagulation: Bleeding and Thrombotic Manifestations of SARS-CoV2 Infection. Blood. 2020;136:489–500.

34. Middeldorp S, Coppens M, van Haaps TF, et al. Incidence of venous thromboembolism in hospitalized patients with COVID-19. J Thromb Haemost. 2020;18:1995–2002.

35. Klok F, Kruip M, Van der Meer N, et al. Incidence of thrombotic complications in critically ill ICU patients with COVID-19. Thromb Res. 2020;191:145–7.

36. Spyropoulos AC. The management of venous thromboembolism in hospitalized patients with COVID-19. Blood adv. 2020;4:4028-.

37. Antithrombotic Therapy in Patients with COVID-19. National Institutes of Health. December 2020. Available at: https://www.covid19treatmentguidelines.nih.gov/adjunctive-therapy/antithrombotic-therapy/. Accessed December 28,2020.

38. Lemos ACB, do Espírito Santo DA, Salvetti MC, et al. Therapeutic versus prophylactic anticoagulation for severe COVID-19: A randomized phase II clinical trial (HESACOVID). Thromb Res. 2020;196:359–66.

39. Gonzalez-Ochoa AJ, Raffetto JD, Hernández AG, et al. Sulodexide in the treatment of patients with early stages of COVID-19: a randomised controlled trial. medrxiv. 2020[E-pub ahead of print];https://doi.org/10.1101/2020.12.04.20242073.

40. NIH ACTIV Trial of blood thinners pauses enrollment of critically ill COVID-19 patients. National Institutes of Health. December 2020. Available at: https://www.nih.gov/news-events/news-releases/nih-activ-trial-blood-thinners-pauses-enrollment-critically-ill-covid-19-patients. Accessed December 24,2020.

41. Barco S, Bingisser R, Colucci G, et al. Enoxaparin for primary thromboprophylaxis in ambulatory patients with coronavirus disease-2019 (the OVID study): a structured summary of a study protocol for a randomized controlled trial. Trials. 2020;21:1–3.

42. Huang C, Wang Y, Li X, et al. Clinical features of patients infected with 2019 novel coronavirus in Wuhan, China. Lancet. 2020;395:497–506.

43. Zhang L, Yan X, Fan Q, et al. D-dimer levels on admission to predict in-hospital mortality in patients with Covid-19. J Thromb Haemost. 2020;18:1324–9.

44. Weinberg I, Fernández-Capitán C, Quintana-Díaz M, et al. Systematic testing for venous thromboembolism in hospitalized patients with COVID-19 and raised D-dimer levels. Thrombosis Update. 2021[E-pub ahead of print];https://doi.org/10.1016/j.tru.2020.100029.

45. Angus DC, Berry S, Lewis RJ, et al. The randomized embedded multifactorial adaptive platform for community-acquired pneumonia (REMAP-CAP) study: rationale and design. Ann Am Thorac Soc. 2020;17:879–91.

46. Normand S-LT. The RECOVERY Platform. N Engl J Med 2020[E-pub ahead of print];https://www.nejm.org/doi/full/10.1056/NEJMe2025674.

47. Ghati N, Roy A, Bhatnagar S, et al. Atorvastatin and Aspirin as Adjuvant Therapy in Patients with SARS-CoV-2 Infection: A structured summary of a study protocol for a randomised controlled trial. Trials. 2020;21:1–3.

48. Vanassche T, Engelen MM, Van Thillo Q, et al. A randomized, open-label, adaptive, proof-of-concept clinical trial of modulation of host thromboinflammatory response in patients with COVID-19: the DAWn-Antico study. Trials. 2020;21:1–14.

49. Houston BL, Lawler PR, Goligher EC, et al. Anti-Thrombotic Therapy to Ameliorate Complications of COVID-19 (ATTACC): Study design and methodology for an international, adaptive Bayesian randomized controlled trial. Clin Trials. 2020;17:491–500.

50. Chen L, Long X, Xu Q, et al. Elevated serum levels of S100A8/A9 and HMGB1 at hospital admission are correlated with inferior clinical outcomes in COVID-19 patients. Cell Mol Immunol. 2020;17:992–4.

51. Lasky JA, Fuloria J, Morrison ME, et al. Design and Rationale of a Randomized, Double-Blind, Placebo-Controlled, Phase 2/3 Study Evaluating Dociparstat in Acute Lung Injury Associated with Severe COVID-19. Adv. Ther. 2020 [E-pub ahead of print];https://doi.org/10.1007/s12325-020-01539-z.

52. Maruyama Y, Yoshida H, Uchino S, et al. Nafamostat mesilate as an anticoagulant during continuous veno-venous hemodialysis: a three-year retrospective cohort study. Int J Artif Organs. 2011;34:571–6.

53. Bikdeli B, Talasaz AH, Rashidi F, et al. Intermediate versus standard-dose prophylactic anticoagulation and statin therapy versus placebo in critically-ill patients with COVID-19: Rationale and design of the INSPIRATION/INSPIRATION-S studies. Thromb Res. 2020;196:382–94.

54. Bikdeli B. Anticoagulation in COVID-19: Randomized trials should set the balance between excitement and evidence. Thromb Res. 2020;196:638–40.

55. Kharma N, Roehrig S, Shible AA, et al. Anticoagulation in critically ill patients on mechanical ventilation suffering from COVID-19 disease, The ANTI-CO trial: A structured summary of a study protocol for a randomised controlled trial. Trials. 2020;21:1–2.

56. Moore HB, Barrett CD, Moore EE, et al. STudy of Alteplase for Respiratory failure in SARS-Cov2/COVID-19: Study Design of the Phase IIa STARS Trial. Res Prac Thromb Haemost. 2020;4:984–96.

57. Dahlberg J, Eriksen C, Robertsen A, Beitland S. Barriers and challenges in the process of including critically ill patients in clinical studies. Scand J Trauma, Resusc Emerg Med. 2020;28:1-8.

58. Schulman S, Kearon C, Scientific SoCoAot, Thrombosis SCotISo, Haemostasis. Definition of major bleeding in clinical investigations of antihemostatic medicinal products in non-surgical patients. J Thromb Haemost. 2005;3:692–4.

59. Mehran R, Rao SV, Bhatt DL, et al. Standardized bleeding definitions for cardiovascular clinical trials: a consensus report from the Bleeding Academic Research Consortium. Circulation. 2011;123:2736–47.

60. Paranjpe I, Fuster V, Lala A, et al. Association of treatment dose anticoagulation with in-hospital survival among hospitalized patients with COVID-19. J Am Coll Cardiol. 2020;76:122–4.

61. Siegal DM, Barnes GD, Langlois NJ, et al. A toolkit for the collection of thrombosis-related data elements in COVID-19 clinical studies. Blood Adv. 2020;4:6259–73.

62. Tritschler T, Mathieu ME, Skeith L, et al. Anticoagulant interventions in hospitalized patients with COVID-19: A scoping review of randomized controlled trials and call for international collaboration. J Thromb Haemost. 2020;18:2958–67.

63. Lopes RD, Fanaroff AC. Anticoagulation in COVID-19: It Is Time for High-Quality Evidence. J Am Coll Cardiol. 2020;76:1827–9.

64. Mousavi S, Moradi M, Khorshidahmad T, Motamedi M. Anti-Inflammatory Effects of Heparin and Its Derivatives: A Systematic Review. Adv Pharmacol Sci. 2015;2015:507151.

65. Bikdeli B, Madhavan MV, Gupta A, et al. Pharmacological agents targeting thromboinflammation in COVID-19: review and implications for future research. Thromb Haemost. 2020;120:1004–24.

66. Deshpande C. Thromboembolic Findings in COVID-19 Autopsies: Pulmonary Thrombosis or Embolism? Ann Intern Med. 2020;173:394–5.

67. McGonagle D, O’Donnell JS, Sharif K, Emery P, Bridgewood C. Immune mechanisms of pulmonary intravascular coagulopathy in COVID-19 pneumonia. Lancet 2020;2:e437–e45.

68. Lawlor M, Gupta A, Ranard LS, et al. Discordance In Activated Partial Thromboplastin Time and Anti-factor Xa Levels in COVID-19 Patients on Heparin Therapy. Thromb Res. 2020;198:79–82.

69. Schünemann HJ, Cushman M, Burnett AE, et al. American Society of Hematology 2018 guidelines for management of venous thromboembolism: prophylaxis for hospitalized and nonhospitalized medical patients. Blood Adv. 2018;2:3198–225.

70. Selvaraj S, Greene SJ, Khatana SAM, Nathan AS, Solomon SD, Bhatt DL. The Landscape of Cardiovascular Clinical Trials in the United States Initiated Before and During COVID-19. J Am Heart Assoc. 2020;9:e018274.

71. Kimmel SE, Califf RM, Dean NE, Goodman SN, Ogburn EL. COVID-19 Clinical Trials: A Teachable Moment for Improving Our Research Infrastructure and Relevance. Ann Intern Med. 2020;173:652–4.

72. Tuttle KR. Impact of the COVID-19 pandemic on clinical research. Nat Rev Nephrol. 2020;16:562–4.

73. Bagiella E, Bhatt DL, Gaudino M. The consequences of the COVID-19 pandemic on non-COVID-19 clinical trials. J Am Coll Cardiol. 2020;76:342–5.

74. Makris M. Staying updated on COVID-19: Social media to amplify science in thrombosis and hemostasis. Res Prac Thromb Haemost. 2020;4:722–6.

75. Angus DC, Alexander BM, Berry S, et al. Adaptive platform trials: definition, design, conduct and reporting considerations. Nat Rev Drug Discov. 2019;18:797–808.

76. Park JJ, Siden E, Zoratti MJ, et al. Systematic review of basket trials, umbrella trials, and platform trials: a landscape analysis of master protocols. Trials. 2019;20:1–10.

77. Horby P, Lim WS, Emberson JR, et al. Dexamethasone in hospitalized patients with Covid-19-preliminary report. N Engl J Med. 2020[E-pub ahead of print];https://www.nejm.org/doi/10.1056/NEJMoa2021436.

78. Consortium WST. Repurposed antiviral drugs for COVID-19—interim WHO SOLIDARITY trial results. N Engl J Med. 2020[E-pub ahead of print];https://www.nejm.org/doi/full/10.1056/NEJMoa2023184.

79. Varshney AS, Wang DE, Bhatt AS, et al. Characteristics of Clinical Trials Evaluating Cardiovascular Therapies for Coronavirus Disease 2019 Registered on ClinicalTrials. gov: A Cross Sectional Analysis. Am Heart J. 2020;232:105–15.

80. FDA Guidance on Conduct of Clinical Trials of Medical Products during COVID-19 Public Health Emergency. FDA Guidance Document. September 2020. Available at: https://www.fda.gov/media/136238/download. Accessed October 30,2020.

81. Spitzer E, Ren B, Brugts JJ, et al. Cardiovascular Clinical Trials in a Pandemic: Immediate Implications of Coronavirus Disease 2019. Card Fail Rev. 2020;6:e09.

82. Vaduganathan M, Butler J, Krumholz HM, Itchhaporia D, Stecker EC, Bhatt DL. Regulation of Cardiovascular Therapies During the COVID-19 Public Health Emergency. J Am Coll Cardiol. 2020;76:2517–21.

83. Gaba P, Bhatt DL. The COVID-19 pandemic: a catalyst to improve clinical trials. Nat Rev Cardiol. 2020;17:673–5.

84. Retracted coronavirus (COVID-19) papers 2020. Available at: https://retractionwatch.com/retracted-coronavirus-covid-19-papers/. Accessed December 28,2020.

85. Pundi K, Perino AC, Harrington RA, Krumholz HM, Turakhia MP. Characteristics and strength of evidence of COVID-19 studies registered on ClinicalTrials. gov “letter”. JAMA Intern Med. 2020;180:1398–400.

86. Dean NE, Gsell P-S, Brookmeyer R, et al. Creating a framework for conducting randomized clinical trials during disease outbreaks. N Engl J Med. 2020;382:1366–9.

87. Sarabipour S, Debat HJ, Emmott E, Burgess SJ, Schwessinger B, Hensel Z. On the value of preprints: An early career researcher perspective. PLoS Biol. 2019;17:e3000151.

